# Symptom Prediction and Mortality Risk Calculation for COVID-19 Using Machine Learning

**DOI:** 10.1101/2021.02.04.21251143

**Authors:** Elham Jamshidi, Amirhossein Asgary, Nader Tavakoli, Alireza Zali, Farzaneh Dastan, Amir Daaee, Mohammadtaghi Badakhshan, Hadi Esmaily, Seyed Hamid Jamaldini, Saeid Safari, Ehsan Bastanhagh, Ali Maher, Amirhesam Babajani, Maryam Mehrazi, Mohammad Ali Sendani Kashi, Masoud Jamshidi, Mohammad Hassan Sendani, Sahand Jamal Rahi, Nahal Mansouri

## Abstract

**Background:** Early prediction of symptoms and mortality risks for COVID-19 patients would improve healthcare outcomes, allow for the appropriate distribution of healthcare resources, reduce healthcare costs, aid in vaccine prioritization and self-isolation strategies, and thus reduce the prevalence of the disease. Such publicly accessible prediction models are lacking, however.

**Methods:** Based on a comprehensive evaluation of existing machine learning (ML) methods, we created two models based solely on the age, gender, and medical histories of 23,749 hospital-confirmed COVID-19 patients from February to September 2020: a symptom prediction model (SPM) and a mortality prediction model (MPM). The SPM predicts 12 symptom groups for each patient: respiratory distress, consciousness disorders, chest pain, paresis or paralysis, cough, fever or chill, gastrointestinal symptoms, sore throat, headache, vertigo, loss of smell or taste, and muscular pain or fatigue. The MPM predicts the death of COVID-19-positive individuals.

**Results:** The SPM yielded ROC-AUCs of 0.53-0.78 for symptoms. The most accurate prediction was for consciousness disorders at a sensitivity of 74% and a specificity of 70%. 2440 deaths were observed in the study population. MPM had a ROC-AUC of 0.79 and could predict mortality with a sensitivity of 75% and a specificity of 70%. About 90% of deaths occurred in the top 21 percentile of risk groups. To allow patients and clinicians to use these models easily, we created a freely accessible online interface at www.aicovid.org.

**Conclusions:** The ML models predict COVID-19-related symptoms and mortality using information that is readily available to patients as well as clinicians. Thus, both can rapidly estimate the severity of the disease, allowing shared and better healthcare decisions with regard to hospitalization, self-isolation strategy, and COVID-19 vaccine prioritization in the coming months.

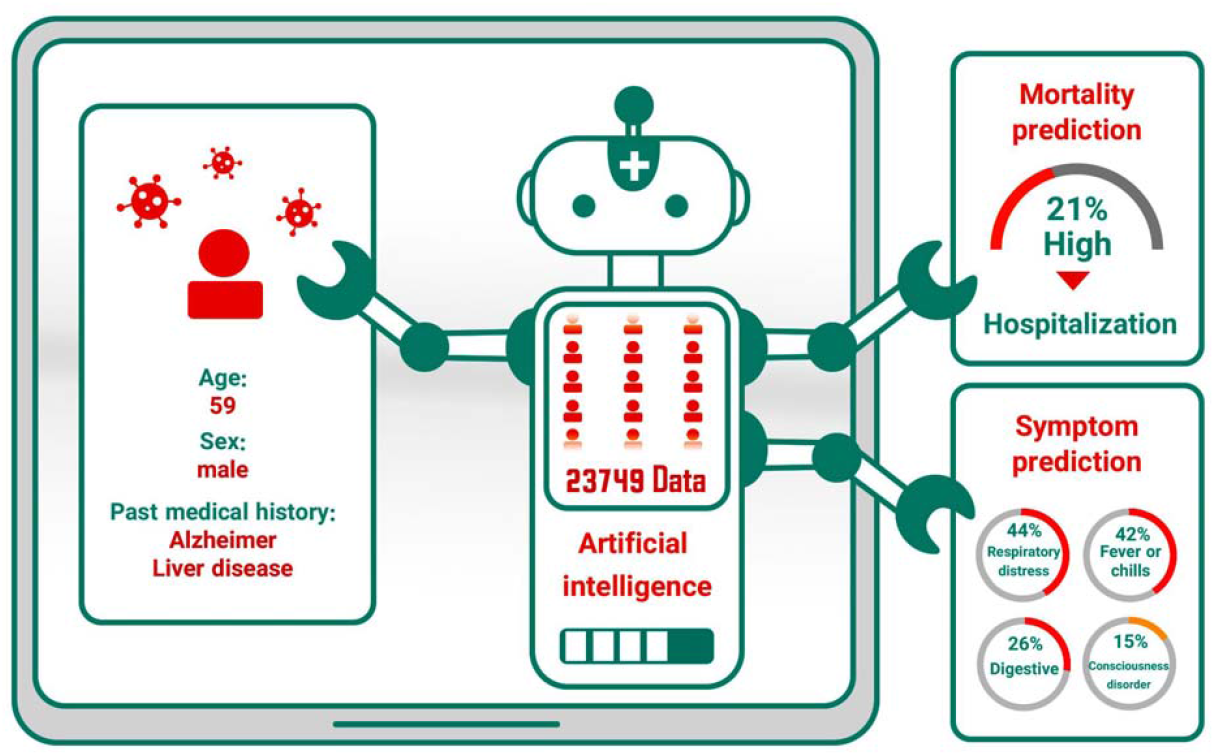

## Introduction

The COVID-19 pandemic of the 2019 novel coronavirus (SARS-CoV-2) started in December 2019 and is spreading rapidly, with approximately 62.5 million confirmed cases and 1.5 million deaths by the end of November 2020.^1^

The severity of the disease varies widely between different patients, ranging from no symptoms, to a mild flu-like illness, to severe respiratory symptoms, and to multi-organ failure leading to death. Among the symptoms, fever, cough, and respiratory distress are more prevalent than symptoms such as consciousness disorders and loss of smell and taste.^2,3^ In general, complications are common among elderly patients and those with pre-existing conditions. The intensive care unit (ICU) admission rate is substantially higher for these groups.^4^

The Center for Disease Control (CDC) and the World Health Organization (WHO) consider the identification of individuals at higher risk a top priority. This identification could be used for numerous solutions to moderate the consequences of the pandemic for the most vulnerable^5^ as well as minimize the presence of actively ill patients in society.

This requires the prediction of the symptoms and mortality risk for infected individuals. While symptom prediction models exist for cancer, no such models have been designed for COVID-19.^6,7^ To make rapid, evidence-based decisions possible, they will ideally be based on readily available patient information, i.e., demographic attributes and past medical history (PMH) as opposed to costly laboratory tests. Early decision making is critical for timely triage and clinical management of patients. For instance, clinical and laboratory data can only be assessed after presenting the individual to a health care center, increasing the risk of unnecessary exposures to the virus and increasing costs.^8^ These parameters are not available immediately and are partly subject to human error. Also, factors like genetic predisposition may increase the models’ accuracy but are not broadly available.

With the growth of big data in healthcare and the introduction of electronic health records, artificial intelligence (AI) algorithms can be integrated into hospital IT systems and have shown promise as computer-aided diagnosis and prognostic tools. In the era of COVID-19, AI has played an essential role in the early diagnosis of infection, the prognosis of hospitalized patients, contact tracing for spread control, and drug discovery.^9^ AI methods can have a higher accuracy over classical statistical analyses.

In contrast to the few previously available COVID-19 risk scales, our mortality prediction model uses a selection of variables that are in principle accessible to all patients and thus can be used immediately after diagnosis.^10,11^ This model not only has a significant benefit in early decision making in the hospital setting, but because it does not require clinicians or laboratories, it can serve as a triage tool for patients in an outpatient setting, in telemedicine, or as a self-assessment tool. For example, decisions on outpatient versus inpatient care can be made remotely by estimating the most probable symptoms and severity risks. This lessens the strain on health care resources, unnecessary costs, and unwanted exposures to infected patients.

Here, we implemented two ML methods to predict the symptoms and the mortality of patients with COVID-19. Overall, 23,749 patients were included in the study. The predictors used for the models were age, sex, and PMH of the patients. Both of these models achieved predictions with high accuracy. To our knowledge, this is one of the largest datasets of COVID-19 cases, and is the only study that uses patient-available data for the prediction of COVID-19 symptoms and mortality. Furthermore, this study is the most extensive study for mortality prediction for COVID-19 using ML based on any set of predictors.^12–15^

We also created an online calculator where each individual can predict their COVID-19 related symptoms and risk (www.aicovid.org).

## Methods

### Source of data and participants

In this cohort study, we used the Hospital Information System (HIS) of 74 secondary and tertiary care hospitals across Tehran, Iran. The eligibility criteria were defined as confirmed or suspected SARS-CoV-2 infections of people aged 18-100 years registered in the referred HIS. The final database used to design the models was obtained by aggregating the 74 hospitals’ HIS. The study included patients referred to any of the hospitals between February 1, 2020, and September 30, 2020. Patients were followed up through October 2020 until all the registered patients had the specific death or survival outcome needed for the mortality prediction model (MPM). This study was approved by the Iran University of Medical Sciences Ethics Committee.

#### Outcome

##### Symptom prediction model

The patients’ symptoms at the time of admission, as recorded in the HIS, were considered as the outputs of the symptom prediction model (SPM). All stated symptoms were clustered in 12 categories to be predicted by the model. The groups are cough, loss of smell or taste, respiratory distress, vertigo, muscular pain or fatigue, sore throat, fever or chill, paresis or paralysis, gastrointestinal problems, headache, chest pain, and consciousness disorders.

##### Mortality prediction model

Death or survival as per the HIS records was defined as the output of the mortality prediction model (MPM).

#### Predictors

The patients’ age, sex, and past medical history (PMH), as detailed in Table 1, were used as predictors for both models. The selection of variables as predictors was based on the available recorded data. All these predictors were recorded in the HIS at the time of admission.

**Table 1.**
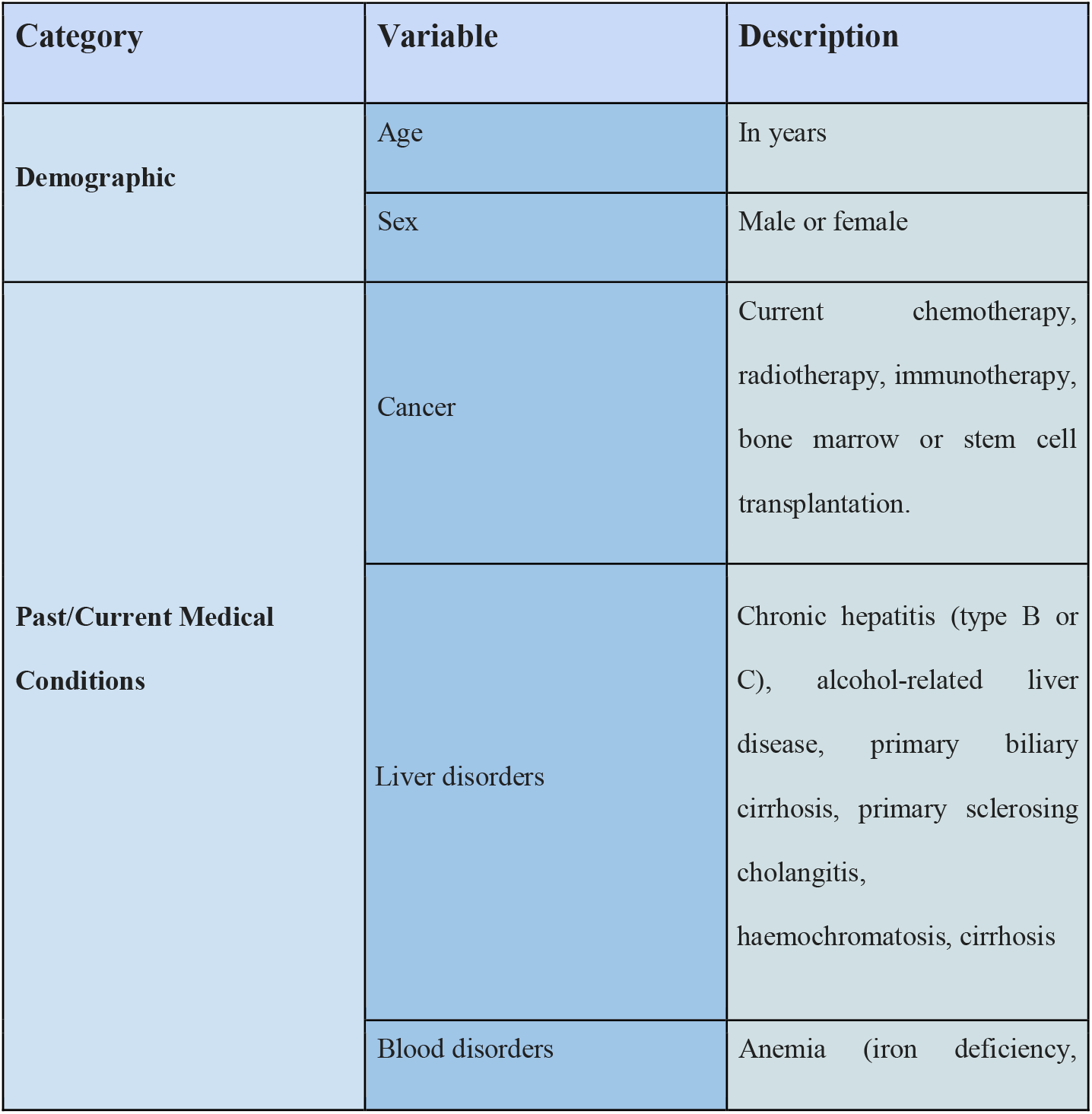
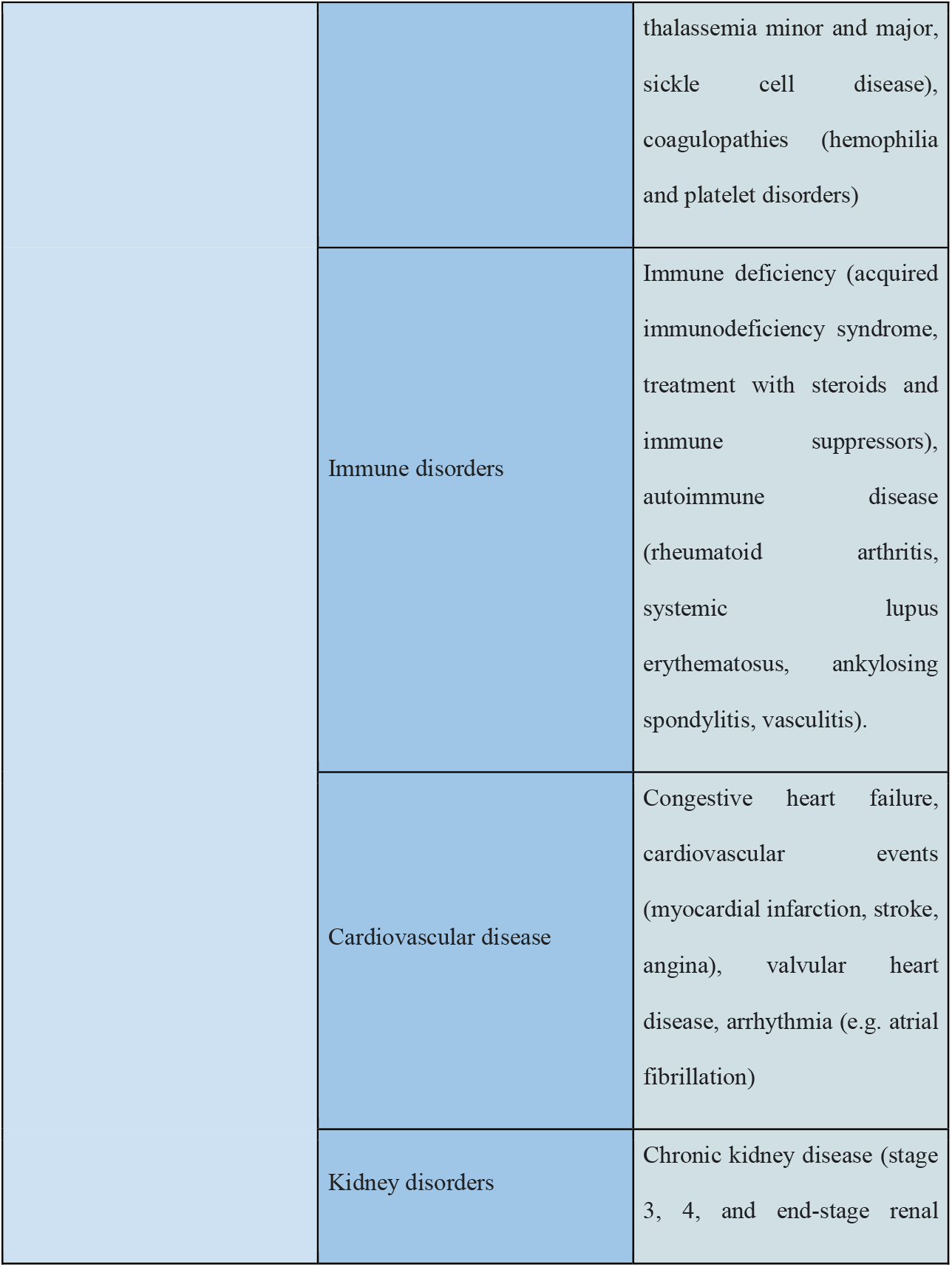
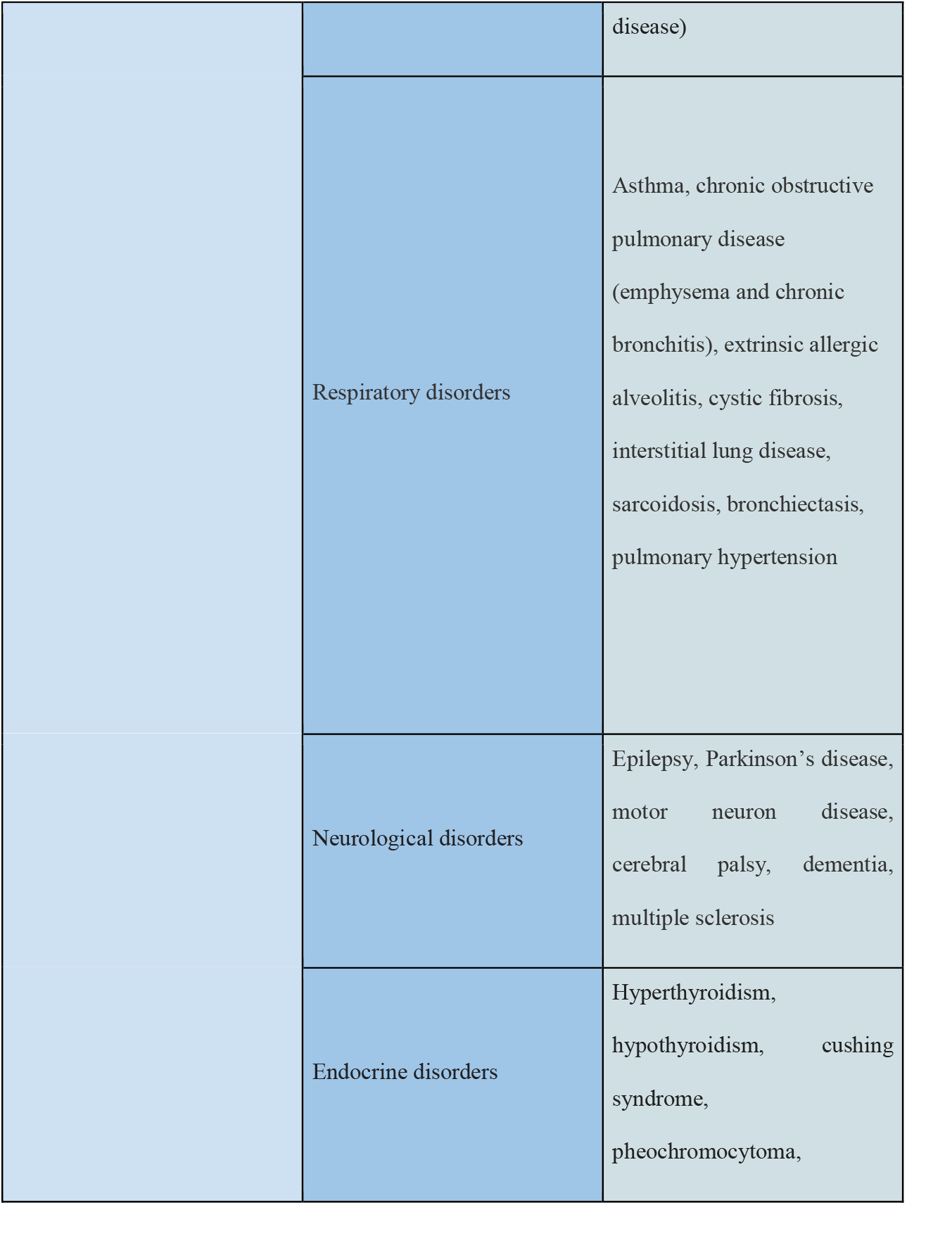
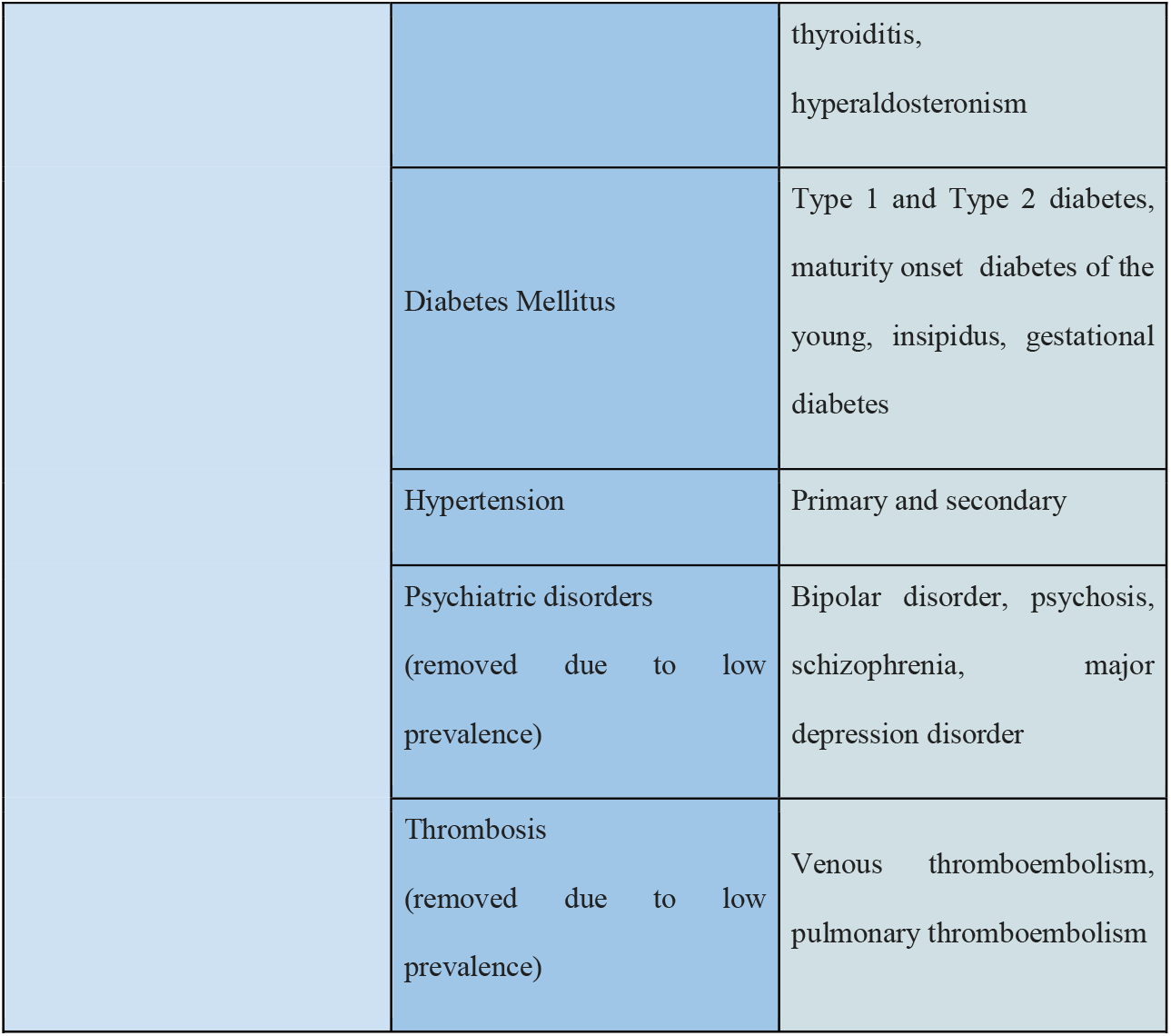
List of predictors. Predictor variables for mortality risk and symptom prediction of COVID-19.

#### Missing data

We only included patients with the required data. Due to the absence of missing data, there was no imputation of missing values.

#### Pre-processing

Symptoms and predictor variables from the medical histories with an incidence of less than 0.2% were removed to reduce noise. This removed past COVID-19 infections, thrombosis, psychiatric disorders, and organ or bone marrow transplantation from the set of predictor variables. The removed symptoms were tachycardia, seizure, nasal congestion, and skin problems.

Sex, PMH, and symptoms were encoded as binary variables. In training and test sets, the only continuous predictor, age, was standardized to zero mean and unit standard deviation.

#### Machine learning methods

To ensure generalizability, a 5-fold cross-validation algorithm was employed.^16^ All records were randomly separated into five independent subsets. Four subsets were used as training data, and one subset was retained as a validation set for model testing. The cross-validation process was then iterated four more times, with each of the five subsets being used as validation data exactly once. Subsequently, model performance metrics were evaluated for training and validation groups separately in each model iteration.

By separating deceased and surviving patients separately into five mortality-stratified subsets first and then combining these into the final five subsets, we maintained the same proportion of deceased and surviving patients in each of the final five subsets.

We evaluated a number of machine learning techniques for both models: Logistic Regression, Random Forest, Artificial Neural Network (ANN), K-Nearest Neighbors (KNN), Linear Discriminant Analysis (LDA), and Naive Bayes.

We calibrated the outputs of the ML models using XYZ.

We took advantage of the Scikit-learn machine learning library to implement both preprocessing algorithms and models.^17^

##### Symptom prediction model

The SPM output predicts symptoms for SARS-CoV-2 positive patients. Since there are 12 symptom groups, we judged the models’ overall performance by a single metric, the prevalence-weighted mean of the twelve ROC-AUCs^18^, in which the ROC-AUCs were weighted by symptom prevalence.

##### Mortality prediction model

The MPM calculates the probability of death for SARS-CoV-2 positive patients. Each model’s performance was measured in terms of a ROC-AUC.

## Results

### Participants

Baseline characteristics of patients and their symptoms are shown in Table 2. Of all 23,749 confirmed or suspected COVID-19 patients, 2,440 (10.27%) passed away at the end of the study (see Discussion). A comparison of the characteristics of survived and deceased patients is shown in Table 3. A comparison of the characteristics of patients with and without each symptom is shown in Appendix Tables 1 to 16.

**Table 2.**
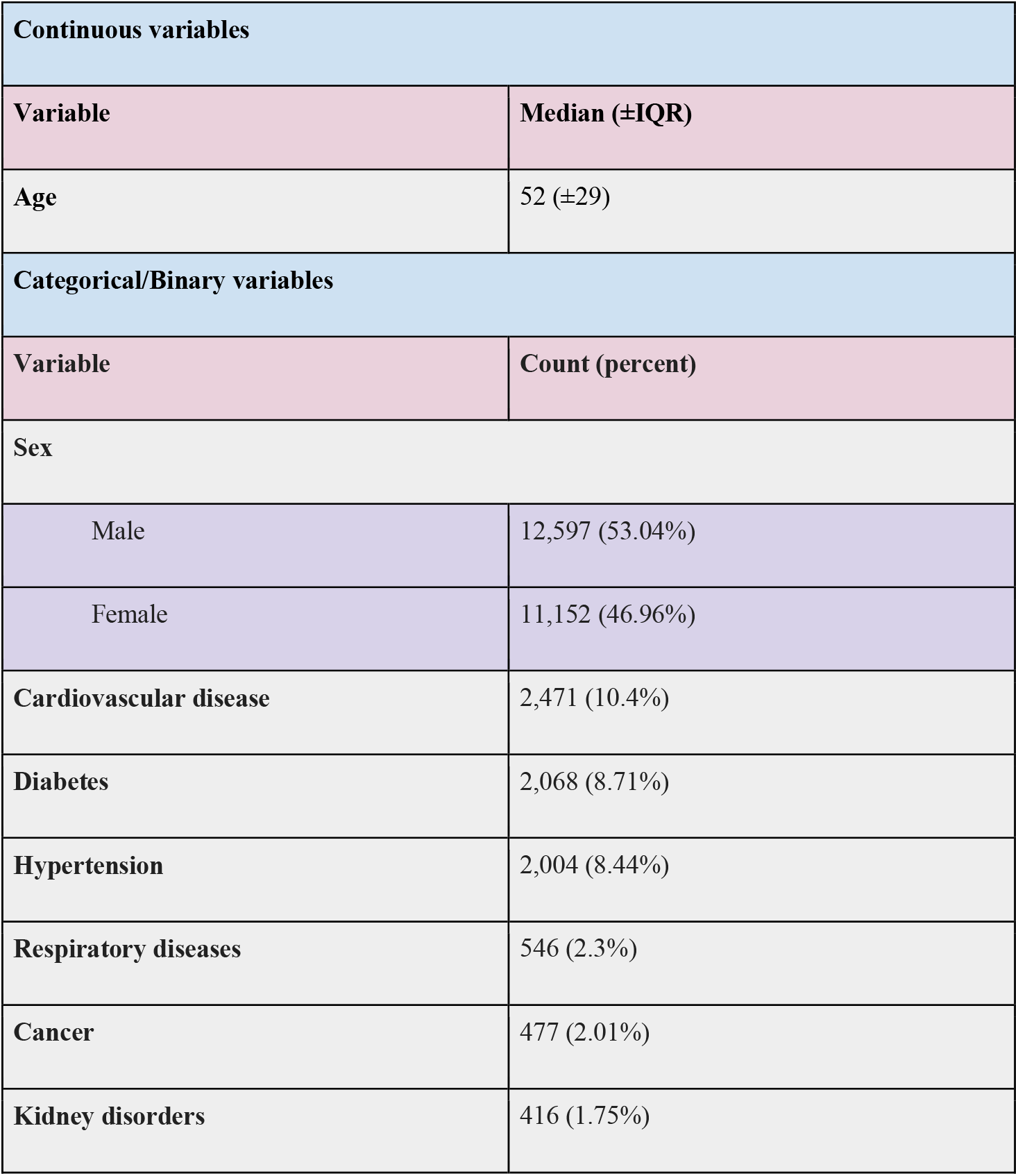
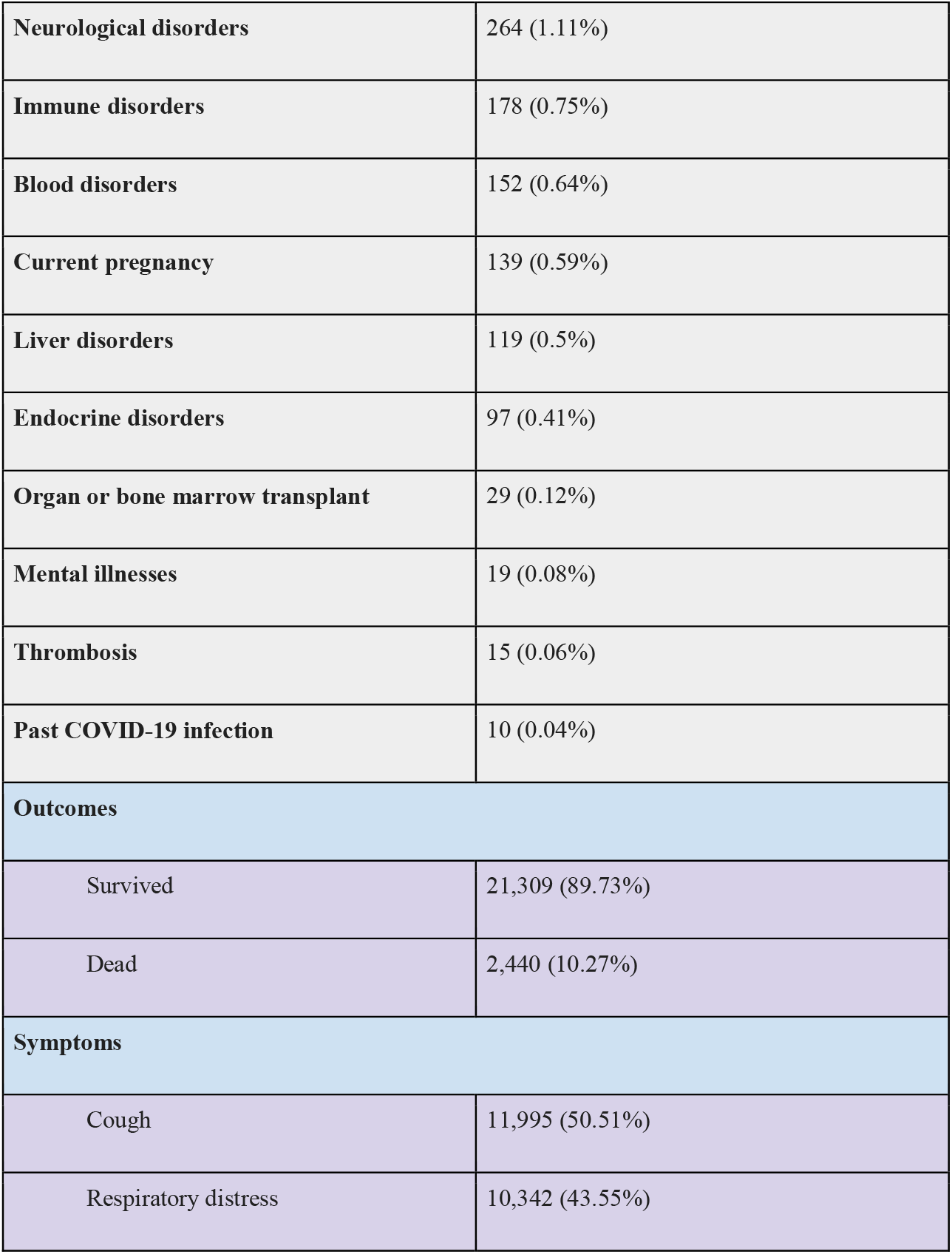
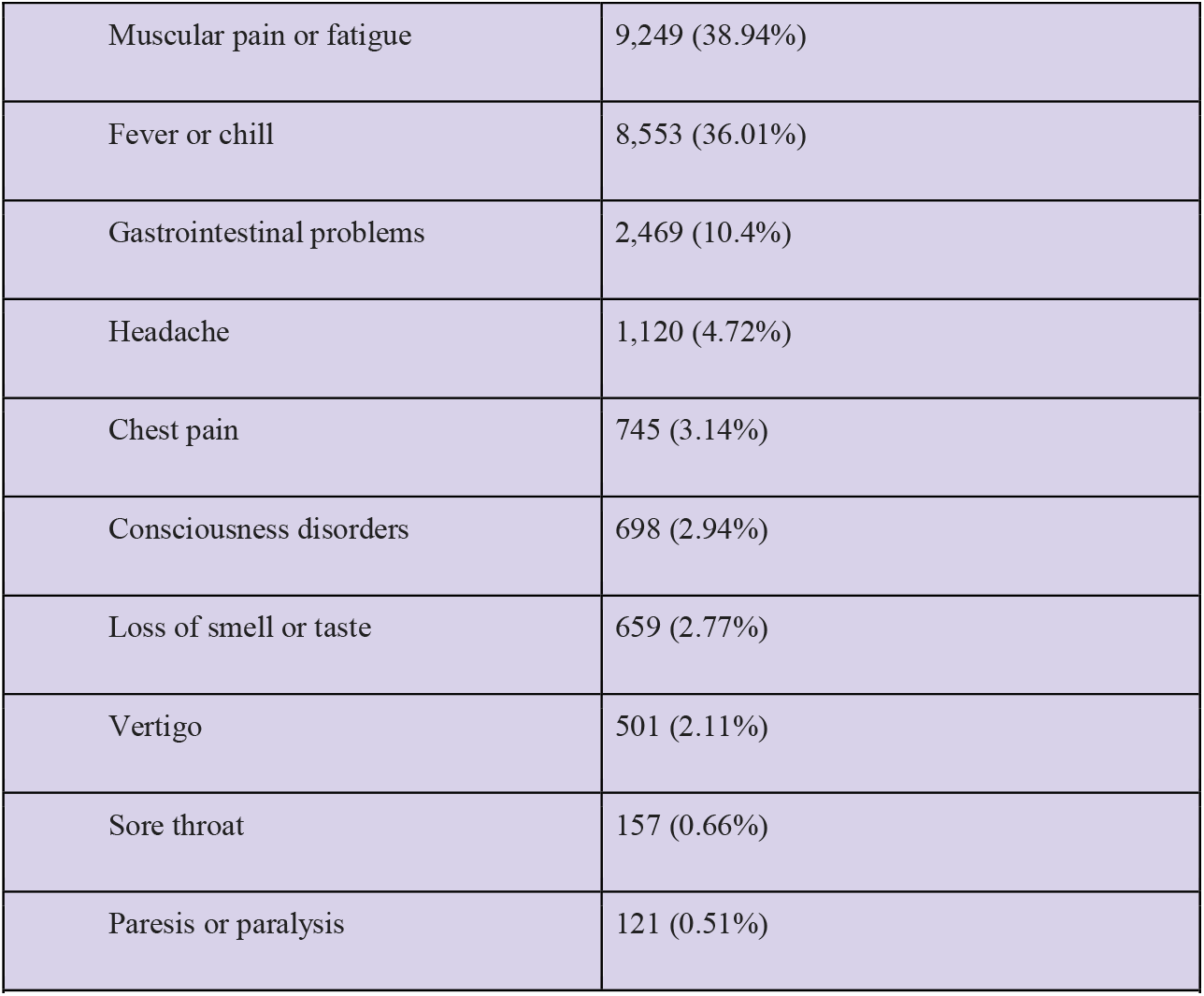
Patient characteristics and symptoms. Baseline characteristics, symptoms, and death outcomes for COVID-19 patients.

**Table 3.** Comparison between survived and deceased patient groups. Comparative evaluation of the characteristics of survived and deceased COVID-19 patients.

| Continuous variables |  |  |  |  |
| --- | --- | --- | --- | --- |
| Variable | Median in survivors (±IQR) | Median in deceased (±IQR) | F-test statistics | F-test p-value |
| Age | 49 (±27) | 70 (±21) | 2,039.47 | <0.001 |
| Categorical/Binary variables |  |  |  |  |
| Variable | Count in survivors (percent in survivors) | Count in deceased (percent in deceased) | Chi2 statistics | Chi2 p-value |
| Sex |  |  |  |  |
| Male | 11,163 (52.39%) | 1,434 (58.77%) | 16.82 | <0.001 |
| Female | 10,146 (47.61%) | 1,006 (41.23%) | 19 | <0.001 |
| Cardiovascular disease | 2,039 (9.57%) | 432 (17.7%) | 139.29 | <0.001 |
| Diabetes | 1,693 (7.94%) | 375 (15.37%) | 138.57 | <0.001 |
| Hypertension | 1,676 (7.87%) | 328 (13.44%) | 80.71 | <0.001 |
| **Respiratory disorders** | 462 (2.17%) | 84 (3.44%) | 15.47 | <0.001 |
| **Cancer** | 343 (1.61%) | 134 (5.49%) | 164.28 | <0.001 |
| **Kidney disorders** | 317 (1.49%) | 99 (4.06%) | 82.54 | <0.001 |
| **Neurological disorders** | 207 (0.97%) | 57 (2.34%) | 36.68 | <0.001 |
| **Immune disorders** | 152 (0.71%) | 26 (1.07%) | 3.62 | 0.057 |
| **Blood disorders** | 112 (0.53%) | 40 (1.64%) | 42.43 | <0.001 |
| **Current pregnancy** | 133 (0.62%) | 6 (0.25%) | 5.35 | 0.021 |
| **Liver disorders** | 101 (0.47%) | 18 (0.74%) | 3.04 | 0.081 |
| **Endocrine disorders** | 88 (0.41%) | 9 (0.37%) | 0.1 | 0.747 |
| **Organ or bone marrow transplant** | 25 (0.12%) | 4 (0.16%) | 0.39 | 0.533 |
| **Psychiatric disorders** | 16 (0.08%) | 3 (0.12%) | 0.63 | 0.428 |
| **Thrombosis** | 13 (0.06%) | 2 (0.08%) | 0.15 | 0.696 |
| **Past COVID-19 infection** | 10 (0.05%) | 0 (0.0%) | 1.15 | 0.285 |

We used statistical hypothesis tests to demonstrate each predictor variable’s significance to the model outputs. We employed the F-test^19^ technique for age, a continuous variable, and the Chi-square^19^ technique for other categorical variables such as sex and PMH.

### Model specification

We evaluated six machine learning methods for both the SPM and MPM, which are listed, together with the hyperparameters used in Table 4.

**Table 4.** Machine learning methods and hyperparameters used.

| The Mortality Prediction Model |  |  |
| --- | --- | --- |
| Method | Parameter | Value(s) |
| Logistic Regression | C | 1.0 |
| Random Forest | Number of trees | 500 |
|  | Min. number of samples at a leaf node | %0.1 of all samples |
|  | Criterion | Gini |
| Artificial Neural Networks | Number of layers | 3 |
|  | Output space dimensionality for each layer | 32, 16, 1 |
|  | Activation function for each layer | Tanh, Tanh, Sigmoid |
| K-Nearest Neighbors | K | 10 |
|  | Weight function | distance |
| Linear Discriminant Analysis | Solver | SVD |
| Naive Bayes | Interval size of age categories | 0.1 |

| The Symptom Prediction Model |  |  |
| --- | --- | --- |
| Method | Parameter | Value |
| Logistic Regression | C | 1.0 |
| Random Forest | Number of trees | 200 |
|  | Min. number of samples at a leaf node | %0.1 of all samples |
|  | Criterion | Gini |
| Artificial Neural Network | Number of layers | 4 |
|  | Output space's dimensionality for each layer | 32, 32, 32, 12 |
|  | Activation function for each layer | Tanh, Tanh, Tanh, Tanh, Sigmoid |
| K-Nearest Neighbors | K | 5 |
|  | Weight function | distance |
| Linear Discriminant Analysis | Solver | SVD |
| Naive Bayes | Interval size of age categories | 0.1 |
| Table 4. Machine learning methods and hyperparameters used. |  |  |

### Model performance

#### Symptom prediction model

The SPM can be considered as 12 separated classifiers; each predicts the occurrence of a specific symptom. While the performance of each sub-classifier can be evaluated separately, the overall performance can be assessed using the prevalence-weighted mean of the ROC-AUCs, since the symptoms have different prevalence. The prevalence-weighted mean ROC-AUC for each method is illustrated in Figure 1. Although the KNN method provided the highest weighted mean ROC-AUC for the test data, it was the least robust method since its performance varied considerably for different validation folds (note standard deviation bars). The Random Forest method achieved better overall performance and robustness. The weighted mean ROC-AUC value of this method was 0.582 for the test data.

**Fig 1.**
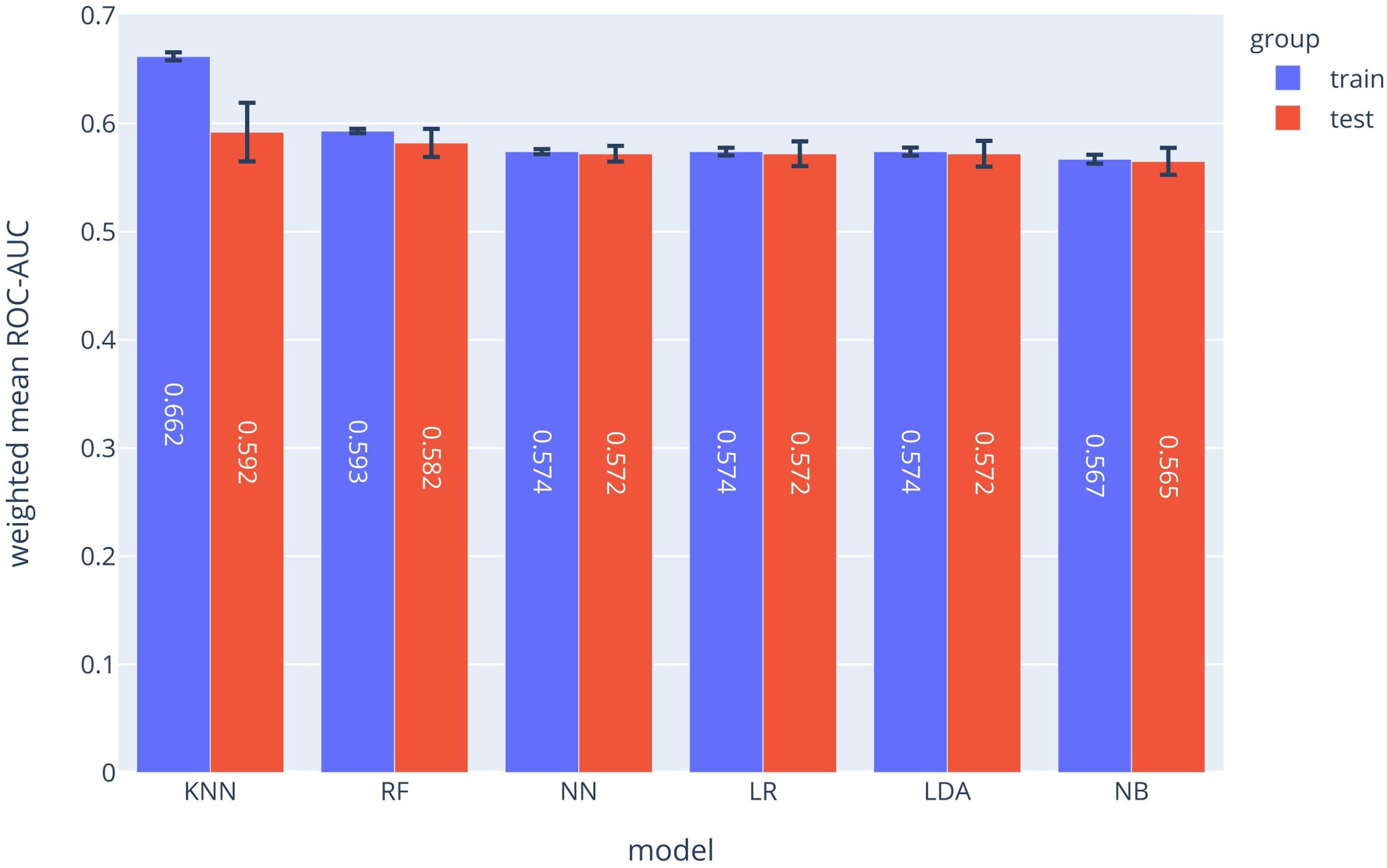
Prevalence-weighted means ROC-AUCs for different ML models. The models were used to implement the Symptom Prediction Model (SPM). Error bars denote the standard deviation over different cross-validation folds.

Moreover, the performance of the SPM can be evaluated for each symptom separately. The ROC-AUC values for predicting consciousness disorder, paresis or paralysis, and chest pain were 0.785, 0.729, and 0.686, respectively. Also, at a specificity of 70%, the sensitivities were 73%, 50%, and 53%, respectively. The comparison between the ROC-AUCs of different ML methods for each symptom is shown in Appendix Figure 1. Appendix Figure 2 shows the performance of each method for all symptoms as a Radar chart. The calibration plot of the RF implementation for each symptom predictor (sub-classifier) is depicted in Appendix Figure 3 which shows the calibration plot of the RF implementation of each symptom.

**Fig 2.**
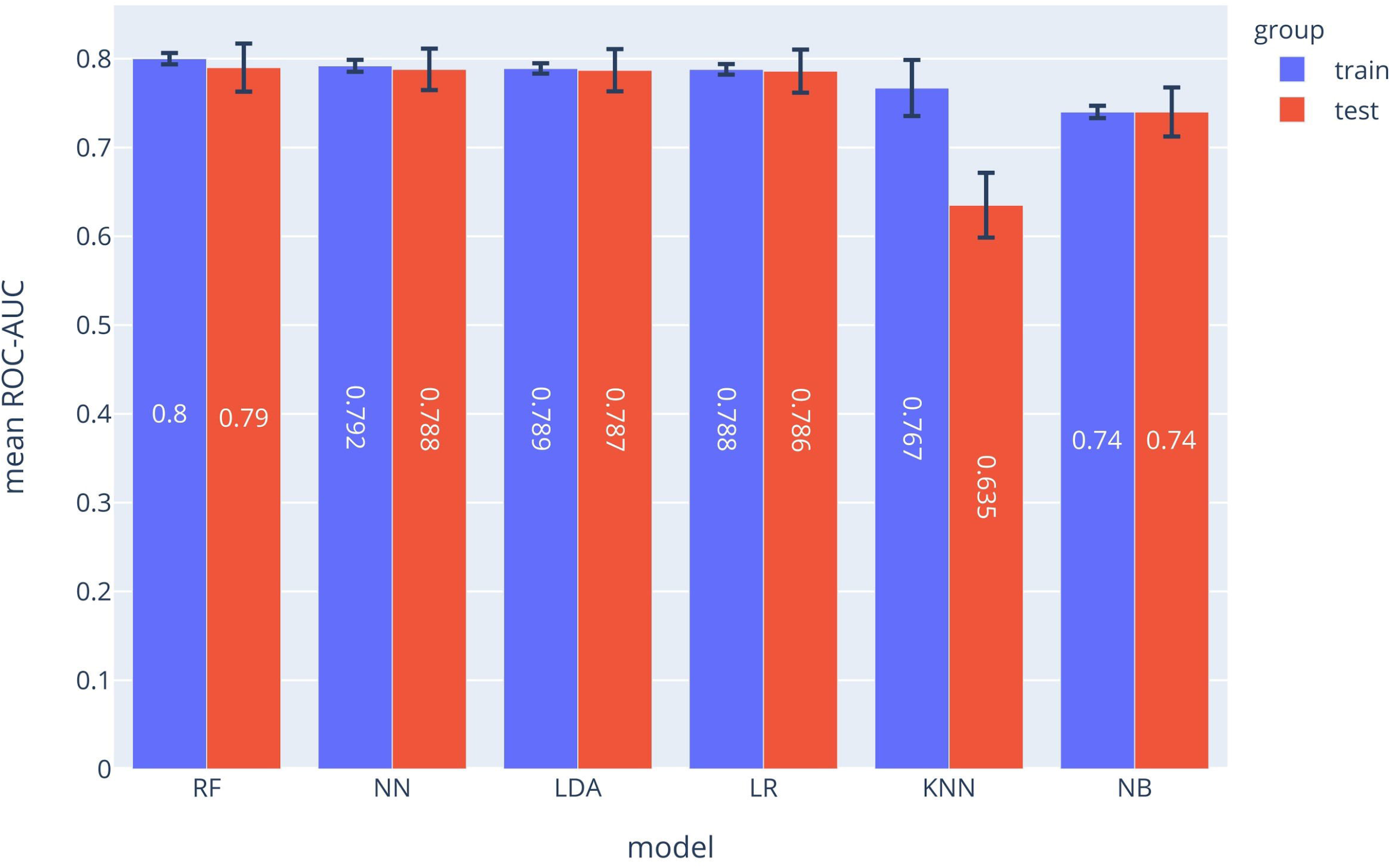
ROC-AUCs of different ML models which were used to implement the MPM. The Random Forest (RF) model outperformed the other approaches.

#### Mortality prediction model

The ROC-AUC values for each method are depicted in Figure 2. In the MPM classifier, the Random Forest method outperformed the other methods just as for the SPM. The achieved ROC-AUC value was 0.79 for the test data.

Appendix Figure 4 shows ROC diagrams representing the true-positive rates versus false-positive rates for each method used to implement the MPM. The calibration plot of the RF model is depicted in Appendix Figure 5.

### Model input-output correlations

We used the Chi-square test and the F-test to evaluate the extent to which PMH, sex, or age predict the outputs of the SPM and MPM. The larger the values of these test values are for each predictor variable, the more the predictor variable is predictive of the output of the models. For categorical predictor variables (i.e., PMH and sex), the Chi-square hypothesis test was used. To evaluate the predictive value of a categorical variable, we examined whether it was more common in patients who died (MPM) or in patients with a particular symptom (SPM). For the only continuous variable (age), we used the F-test. To find the impact of age, we examined if the age median was higher in dead patients (MPM) or in patients with a particular symptom (SPM). For the SPM model, Appendix Figure 6 shows how each factor in the PMH was correlated with each symptom using the Chi-square test. For example, patients with diabetes or cardiovascular disease were more likely to have consciousness disorders and chest pain in case of infection with COVID-19. The effect of age on each symptom is shown in Appendix Figure 7 using the F-test. Older patients were more likely to develop symptoms such as respiratory distress and consciousness disorder but also less likely to develop symptoms such as muscular pain or fatigue.

In addition, for the MPM, the impact of each PMH on death is shown in Appendix Figure 8. In our analysis, cancer, cardiovascular disease, and diabetes have the greatest effects on the risk of death in patients with Covid 19; on the other hand, pregnancy or being female decreased the chances of death. The F-test statistic of age in the MPM model is 2,039.47, which explains the increase in mortality risk from aging.

## Discussion

Our objective in this study was to develop two ML models to predict the mortality and symptoms of COVID-19-positive patients among the general population using age, gender, and comorbidities alone. These models can guide the design of measures to combat the COVID-19 pandemic. The prediction of vulnerability using the models allows people in different risk groups to take appropriate actions if they contract COVID-19. For example, people who fall into the low-risk group can start isolation sooner when the predicted symptoms appear and refer to a hospital only if the symptoms persist. As a result, the risk of disease spread and the pressure on the health care system from unnecessary hospital visits, costs, and psychological and physical stress to the medical staff could be reduced.^20^ In contrast, people who are predicted to be at higher risk are recommended to seek medical care immediately. Predictions can speed up the treatment process and ultimately decrease mortality.

Our study has shown that multiple symptoms have strong correlations with different medical history factors. Symptoms can be either amplified or attenuated by health backgrounds; for instance, hypertension, diabetes, and respiratory and neurological disorders increased the chances of loss of smell or taste; however, pregnancy, cancer, higher age, cardiovascular disease, and liver, immune system, blood, and kidney disorders have attenuated the appearance of this symptom.

Due to the complexity of the COVID-19 pathogenesis, many clinical studies revealed contradictory results, for example, the effectiveness or ineffectiveness of Remdisivir.^21–23^ We hypothesize that the imbalance of mortality risks between the intervention and control groups could have been a problem in these studies. With the help of our model, such problems could be partially solved by equalizing the mortality baseline in different clinical groups.

Our AI models can also be beneficial for COVID-19 vaccine testing and prioritization strategies. The limited number of approved vaccines in the first months of the vaccination process and the potential shortages make vaccine prioritization inevitable. This prioritization would be more important for developing countries which do not have the resources to pre-order vaccines from multiple companies.^24^ Having a mortality prediction tool for each individual could be a valuable tool for governments to decide on vaccines’ allocation.

## Limitations

Since our dataset was collected by the HIS, it did not contain COVID-19 patients that did not refer to a hospital or had no major symptoms to be identified as infected. This could explain the high mortality rate in our and other studies.^15,25,26^ However, for a systematic study with few confounding variables, uniform data collection is essential, which can only be realistically ensured with hospital data.

Another limitation could be that late deaths that occurred after October 2020 were not included in our study.

Also, other variables such as the viral load may be important but are difficult to measure and are not readily available. We opted for easily accessible predictor variables to allow the widespread use of the models.

One way to improve the models is to subgroup specific factors in the medical history or specific symptoms further. The main reason for grouping factors and symptoms was the low prevalence of certain subsets in the dataset.

In conclusion, we evaluated 15 parameters (Table 1) for predicting the symptoms and the mortality risk of COVID-19 patients. The ML models trained in this study could help people quickly determine their mortality risk and the probable symptoms of the infection. These tools could aid patients, physicians, and governments with informed and shared decision making.

## Data Availability

The data that support the findings of this study are available from the corresponding authors upon request.

## Authors Contribution

Elham Jamshidi: Conceptualization-Equal, Methodology-Equal, Project administration-Equal, Writing-original draft-Equal, Writing-review & editing-Equal

Amirhossein Asgary: Conceptualization-Equal, Methodology-Equal, Project administration-Equal, Writing-original draft-Equal, Writing-review & editing-Equal

Nader Tavakoli: Conceptualization-Equal, Methodology-Equal, Project administration-Equal, Writing-original draft-Equal, Writing-review & editing-Equal

Alireza Zali: Data curation-Equal, Investigation-Equal, Writing-original draft-Equal, Writing-review & editing-Equal

Farzaneh Dastan: Data curation-Equal, Investigation-Equal, Writing-original draft-Equal, Writing-review & editing-Equal

Amir Daaee: Data curation-Equal, Investigation-Equal, Writing-original draft-Equal, Writing-review & editing-Equal

Mohammadtaghi Badakhshan: Data curation-Equal, Investigation-Equal, Writing-original draft-Equal, Writing-review & editing-Equal

Hadi Esmaily: Data curation-Equal, Investigation-Equal, Writing-original draft-Equal, Writing-review & editing-Equal

Seyed Hamid Jamaldini: Data curation-Equal, Investigation-Equal, Writing-original draft-Equal, Writing-review & editing-Equal

Saeid Safari: Data curation-Equal, Investigation-Equal, Writing-original draft-Equal, Writing-review & editing-Equal

Ehsan Bastanhagh: Data curation-Equal, Investigation-Equal, Writing-original draft-Equal, Writing-review & editing-Equal

Ali Maher: Data curation-Equal, Investigation-Equal, Writing-original draft-Equal, Writing-review & editing-Equal

Amirhesam Babajani: Data curation-Equal, Investigation-Equal, Writing-original draft-Equal, Writing-review & editing-Equal

Maryam Mehrazi: Data curation-Equal, Investigation-Equal, Writing-original draft-Equal, Writing-review & editing-Equal

Mohammad Ali Sendani Kashi: Data curation-Equal, Investigation-Equal, Writing-original draft-Equal, Writing-review & editing-Equal

Masoud Jamshidi: Data curation-Equal, Investigation-Equal, Writing-original draft-Equal, Writing-review & editing-Equal

Mohammad Hassan Sendani: Data curation-Equal, Investigation-Equal, Writing-original draft-Equal, Writing-review & editing-Equal

Sahand Jamal Rahi: Conceptualization-Equal, Methodology-Equal, Project administration-Equal, Supervision-Equal, Writing-original draft-Equal, Writing-review & editing-Equal

Nahal Mansouri: Conceptualization-Equal, Methodology-Equal, Project administration-Equal, Supervision-Equal, Writing-original draft-Equal, Writing-review & editing-Equal

## Abbreviation list

AI: artificial intelligence
COVID-19: coronavirus disease of 2019
ICU: intensive care unit
IQR: interquartile range
KS: Kolmogorov-Smirnov
LR: logistic regression
ML: machine learning
RF: random forest
RDW: red blood cell distribution width
ROC: receiver operating characteristic
HIS: (hospital information system

## Declaration of competing interest

The authors declare having no potential conflicts of interest.

## Data availability statement

The data that support the findings of this study are available from the corresponding authors upon reasonable request.

## Notes

### Competing Interest Statement

The authors have declared no competing interest.

### Funding Statement

SJR thanks the Ecole polytechnique federale de Lausanne for generous support

### Author Declarations

The study was performed after approval by Iran University of Medical Sciences Ethics Committee

