## Supplementary Material for "Symptom Prediction and Mortality Risk Calculation for COVID-19 Using Machine Learning"

### Appendix Figures

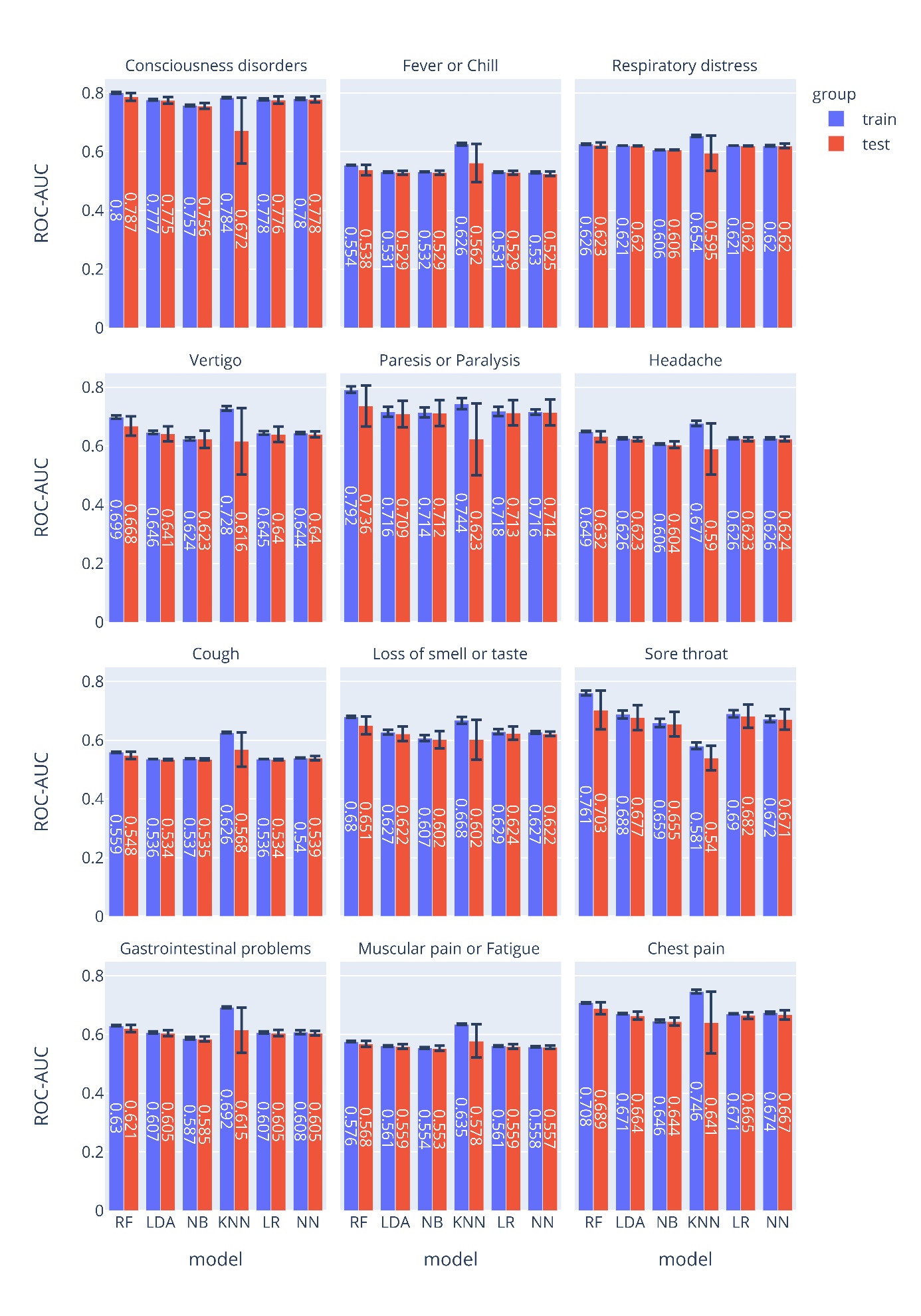

**Appendix Figure 1.**  **ROC-AUCs for specific COVID-19 symptoms**.

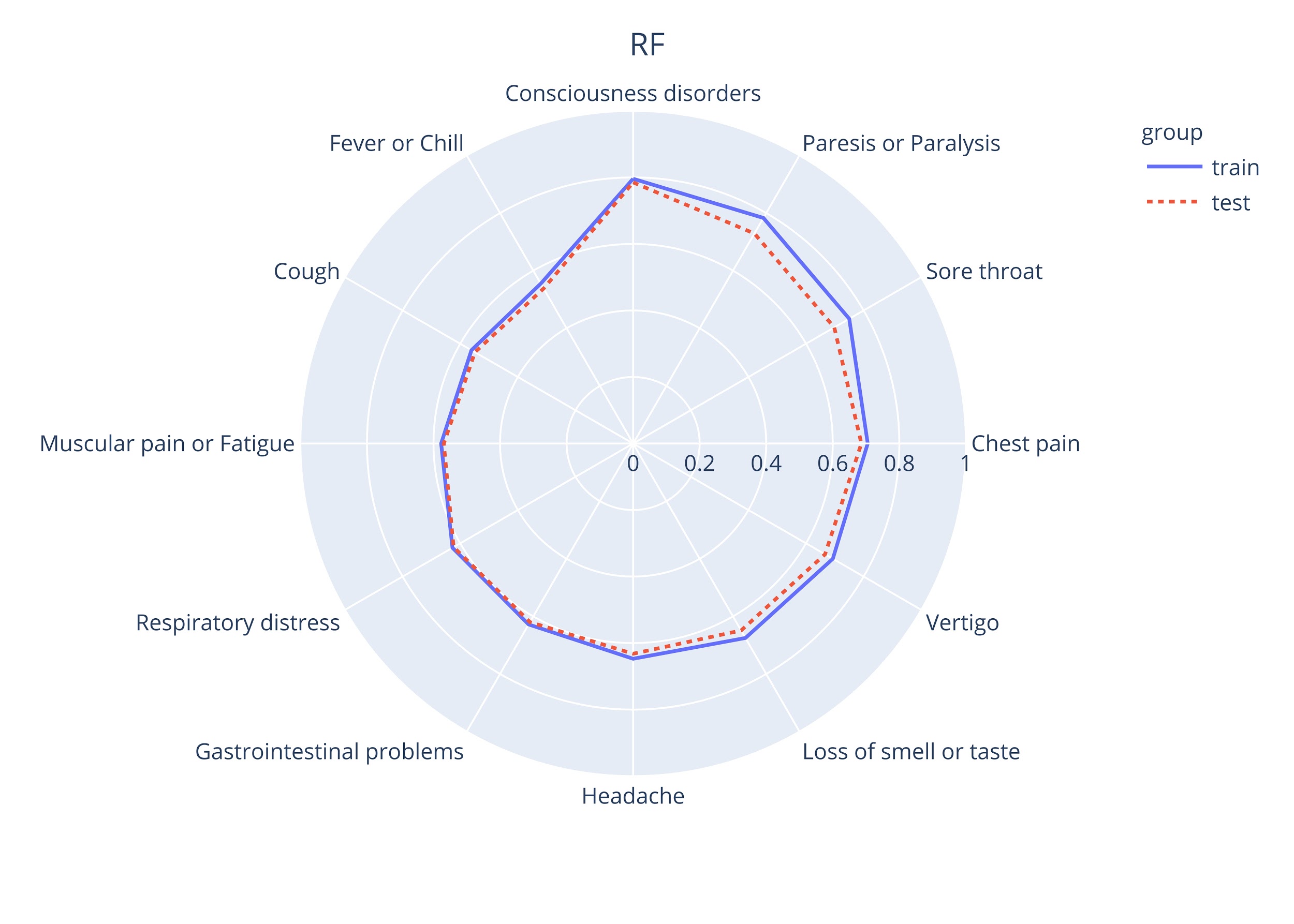

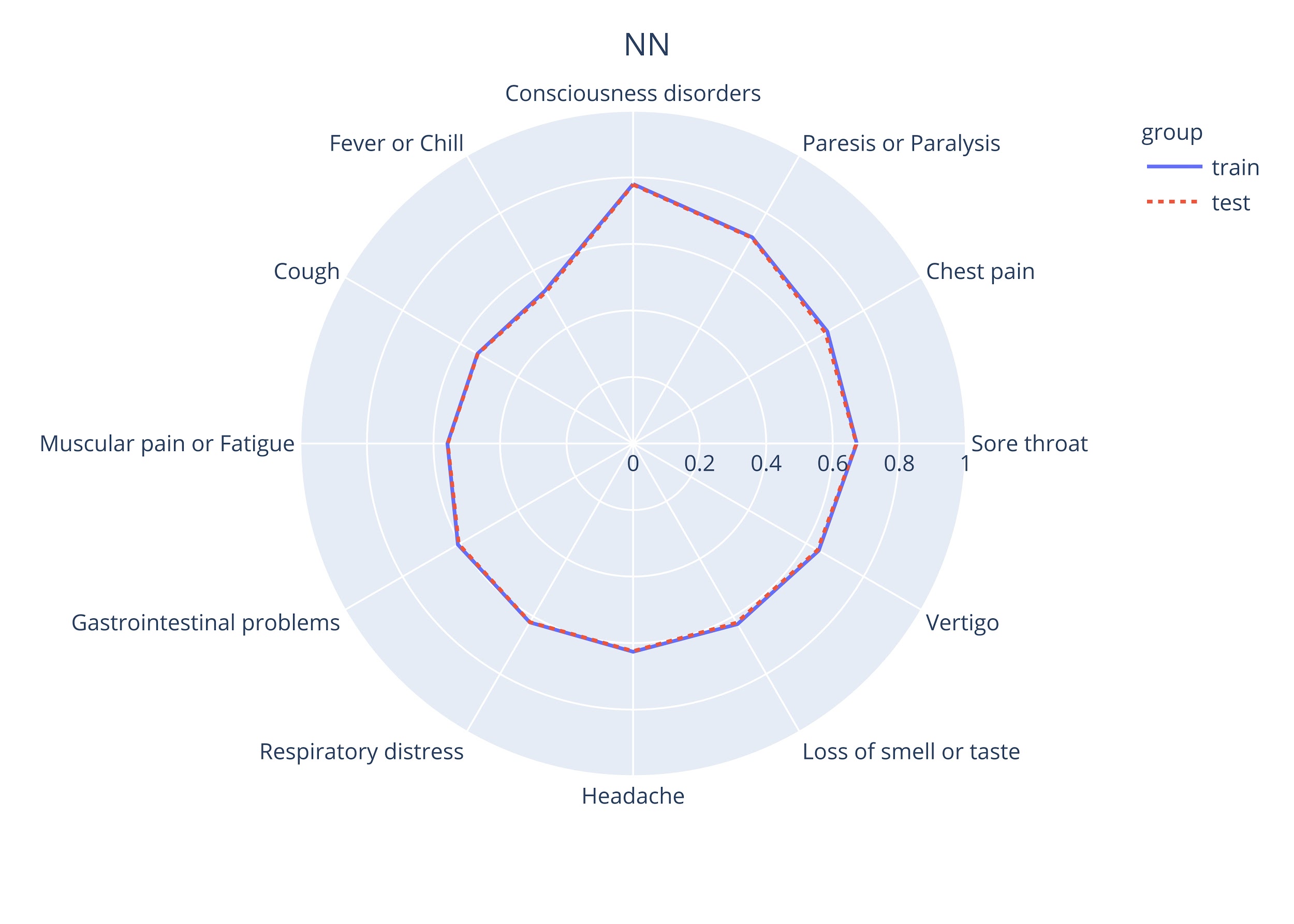

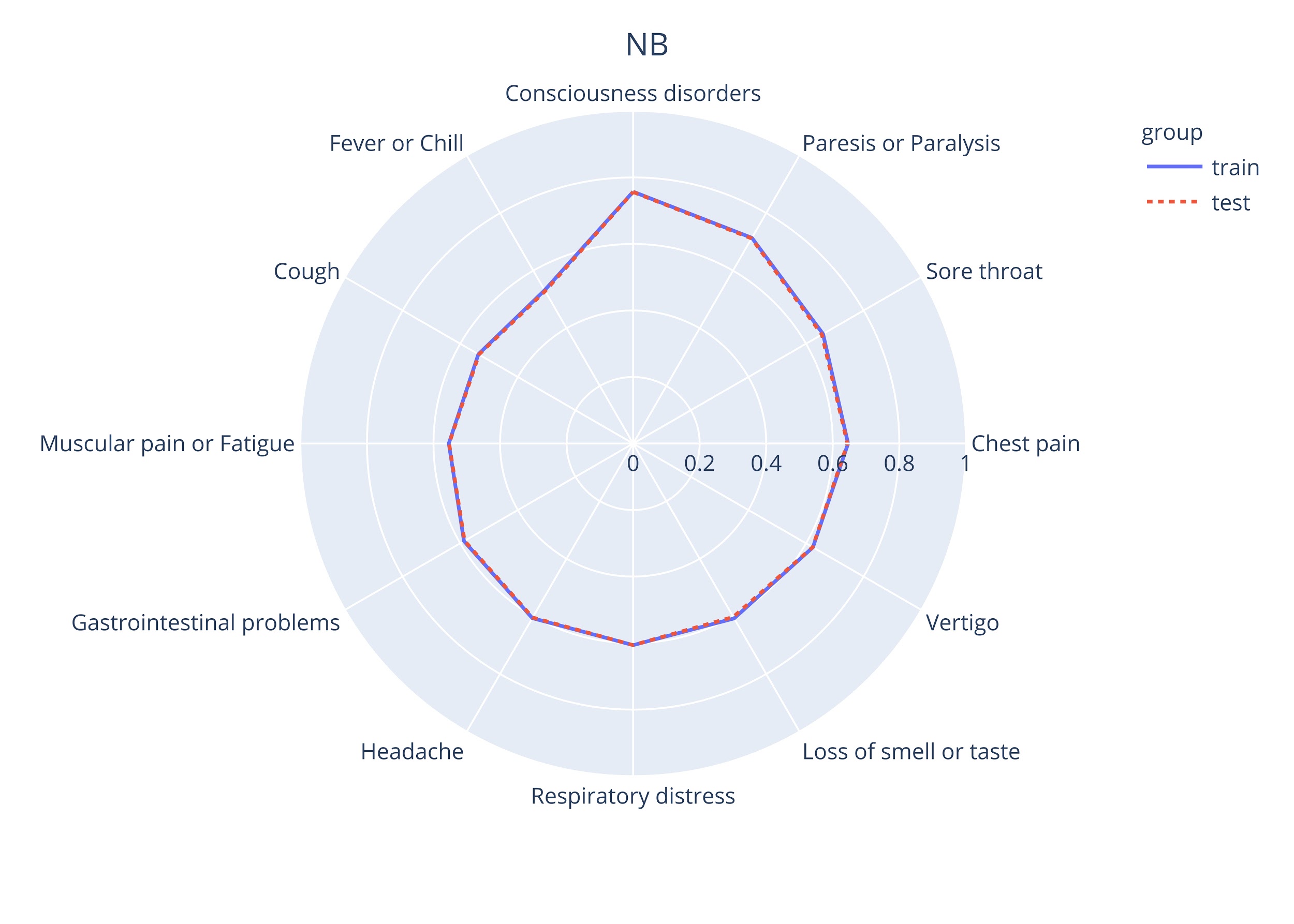

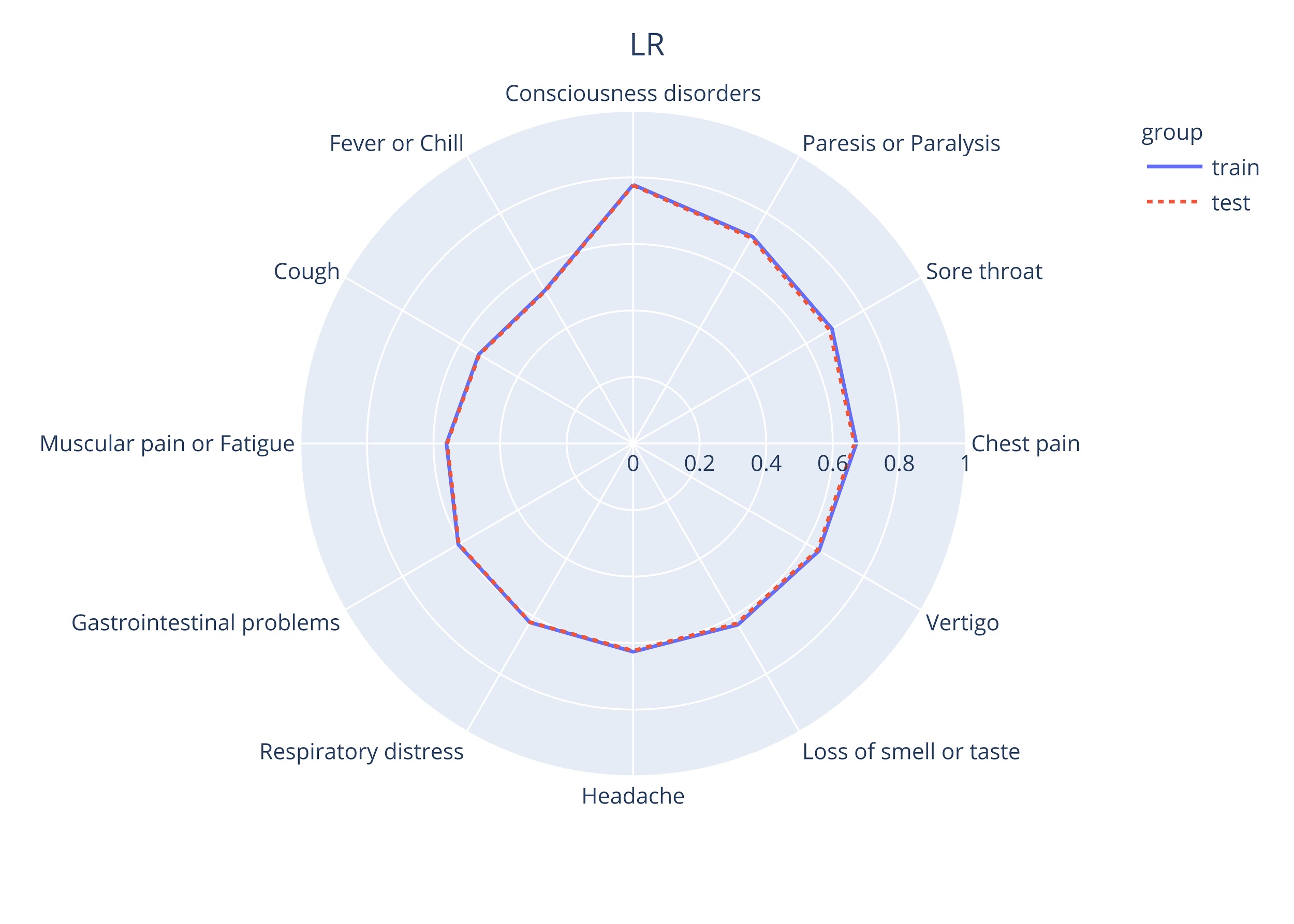

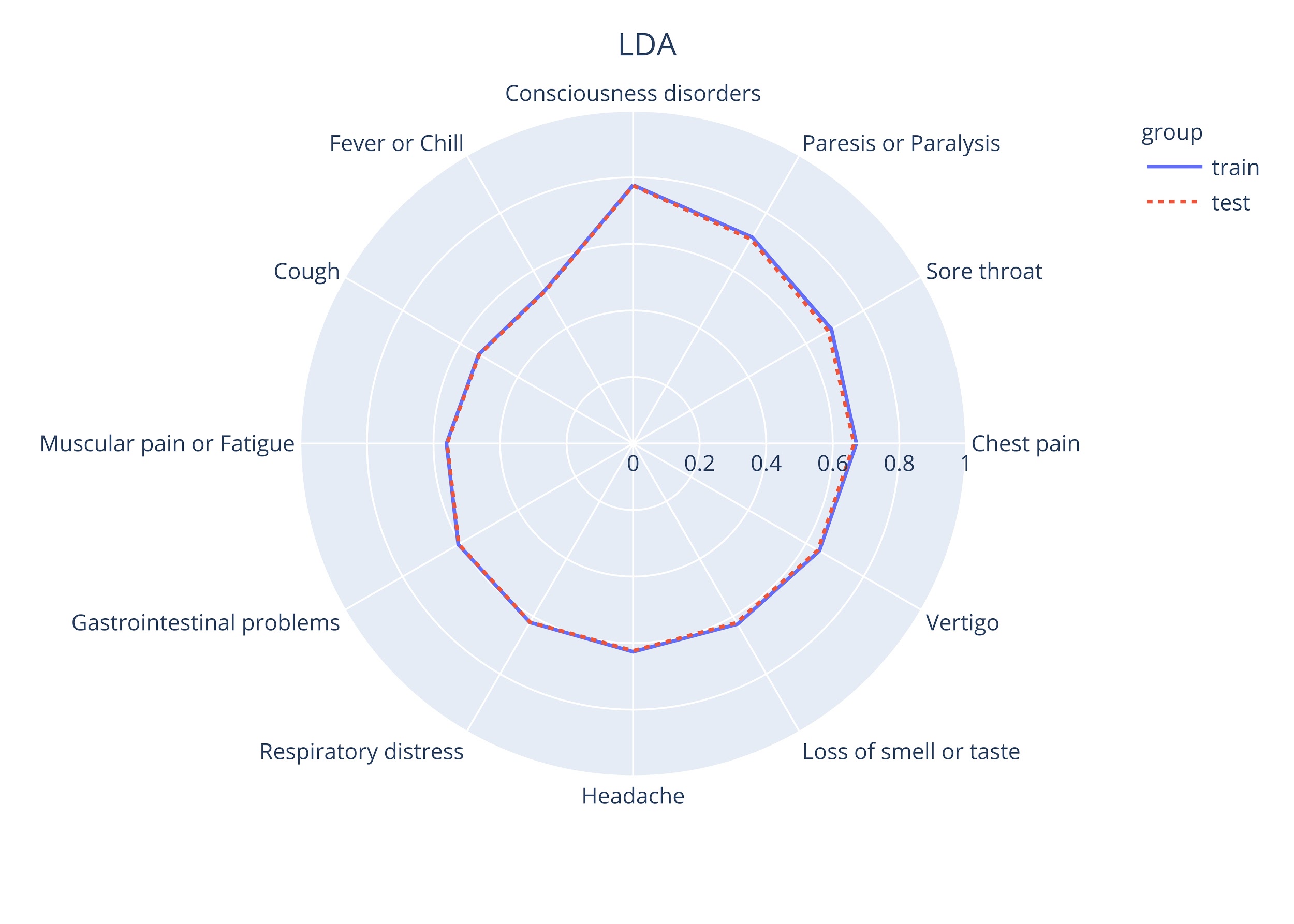

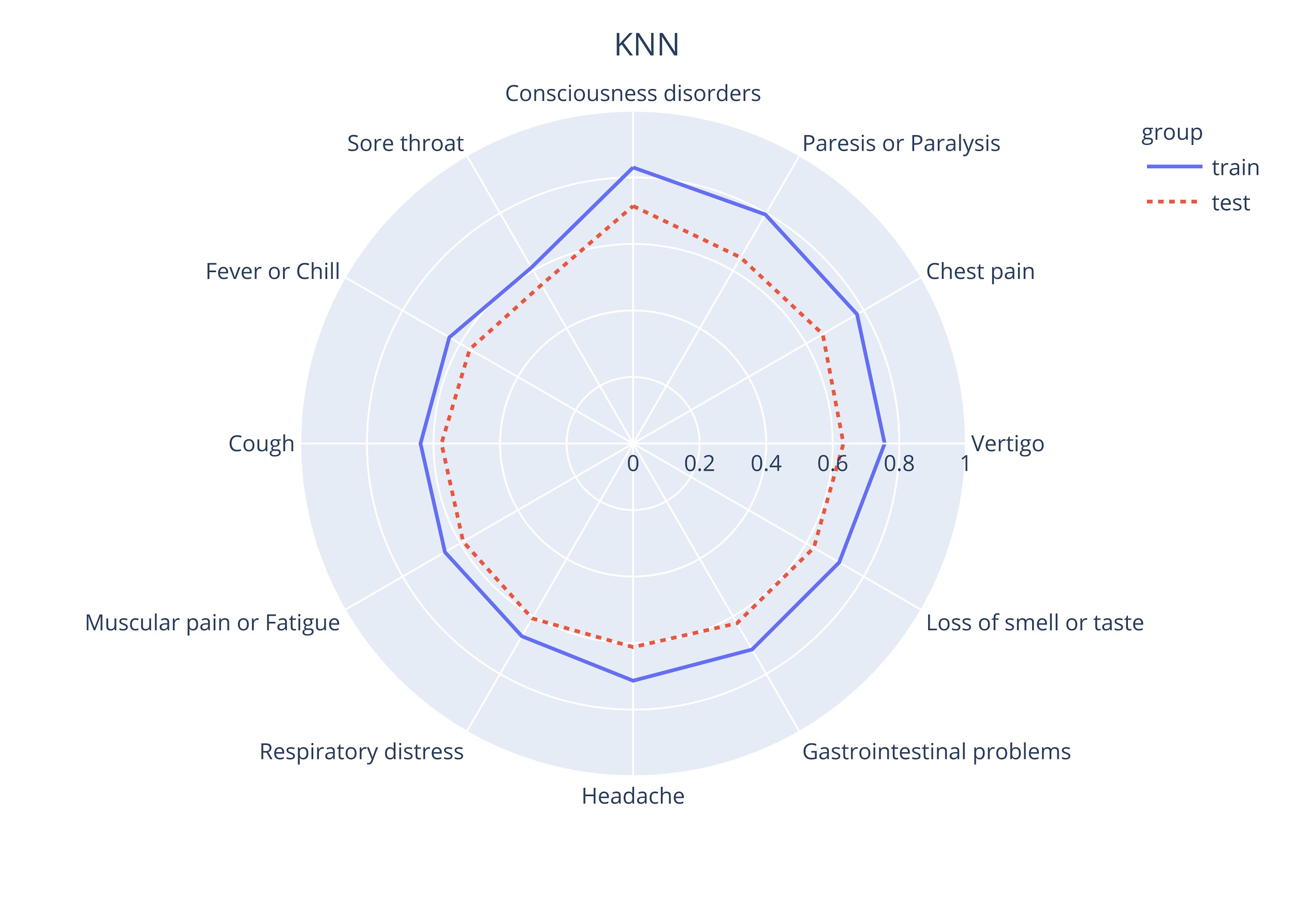

**Appendix Figure 2.** **Performance of ML methods for specific symptoms.**

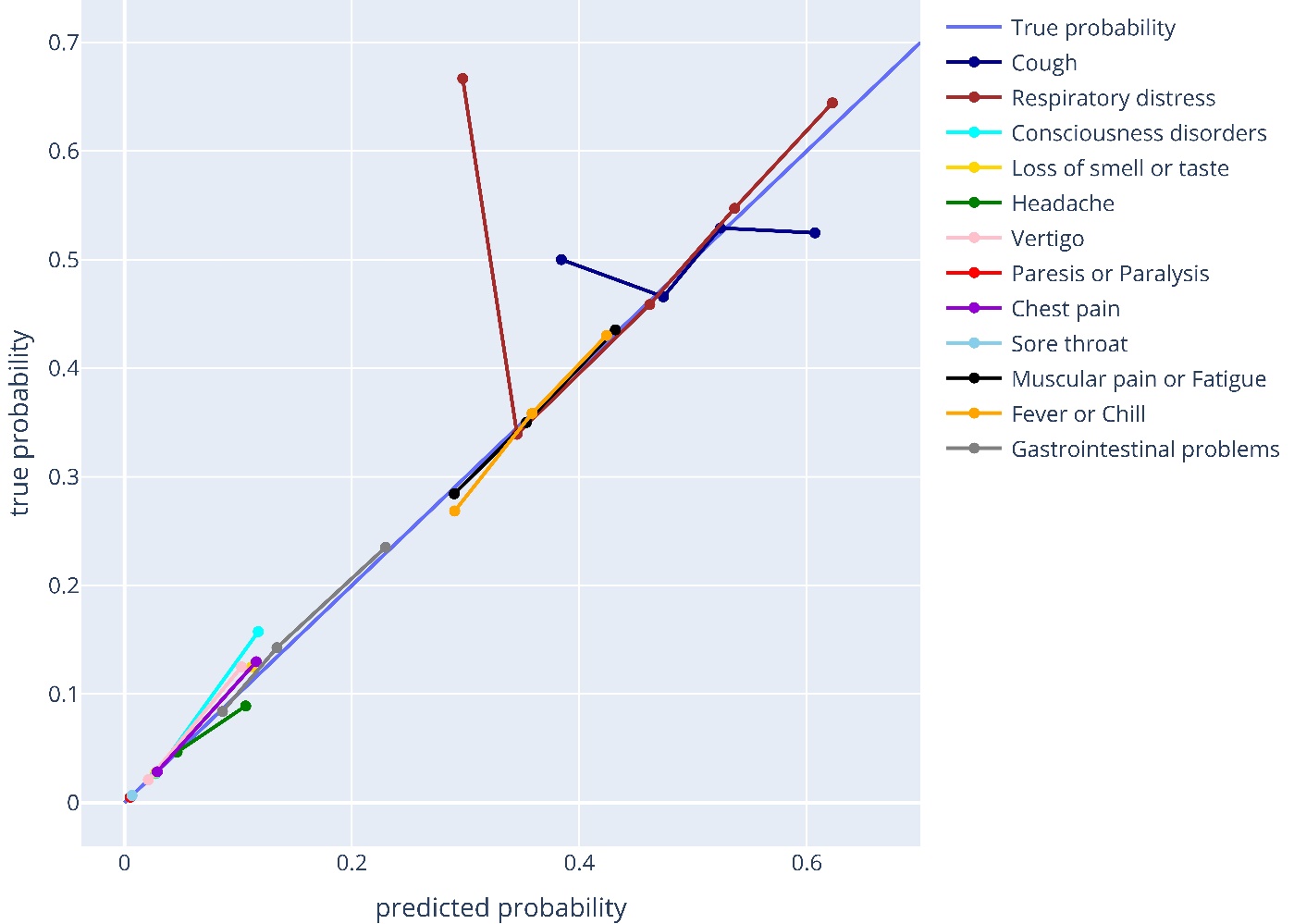

**Appendix Figure 3**. **Calibration of predicted probability symptoms in the SPM.**

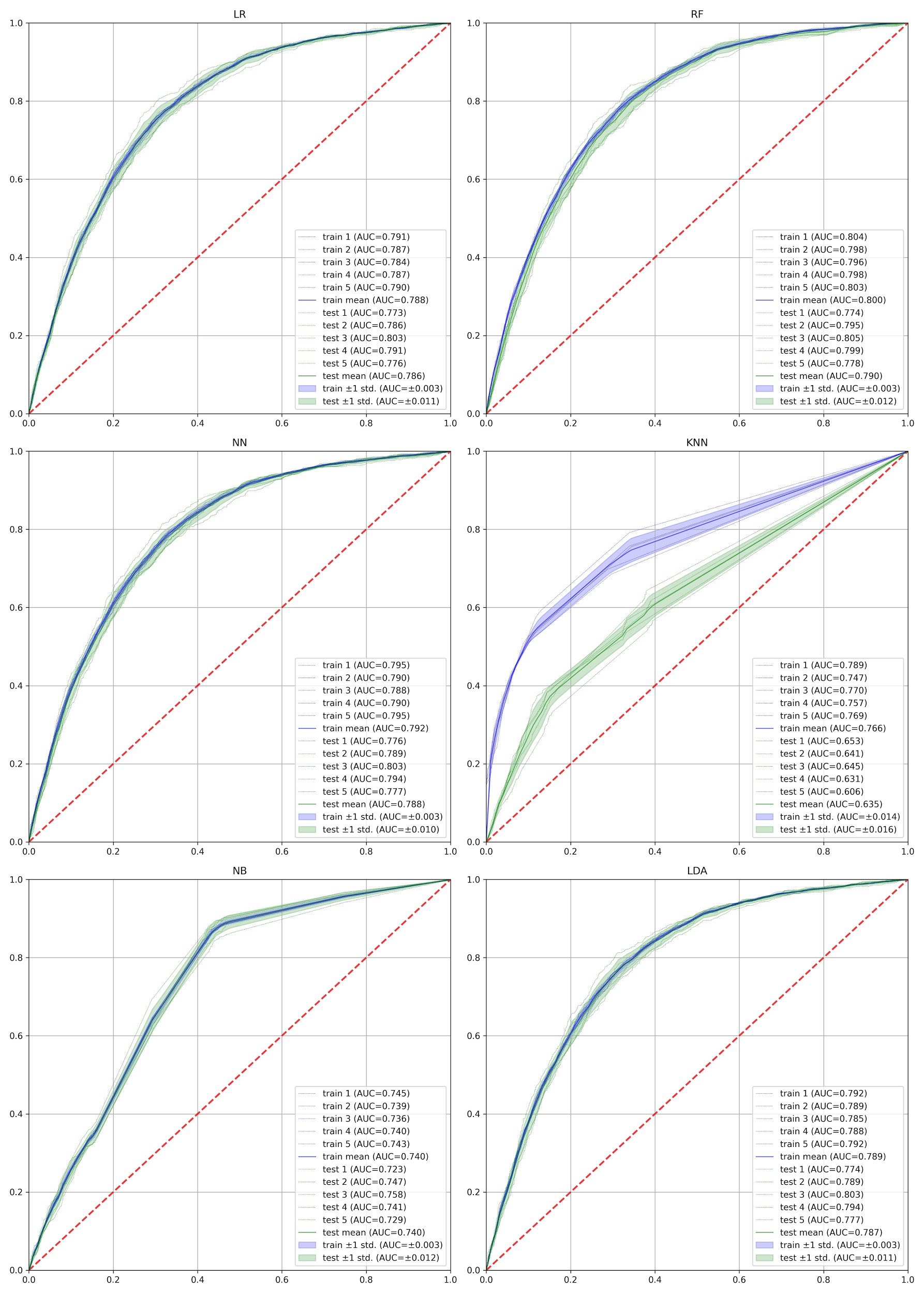

**Appendix Figure 4.** **ROCs of different ML methods which were used to implement the MPM.** The ROC curves of each cross-validation fold are depicted.

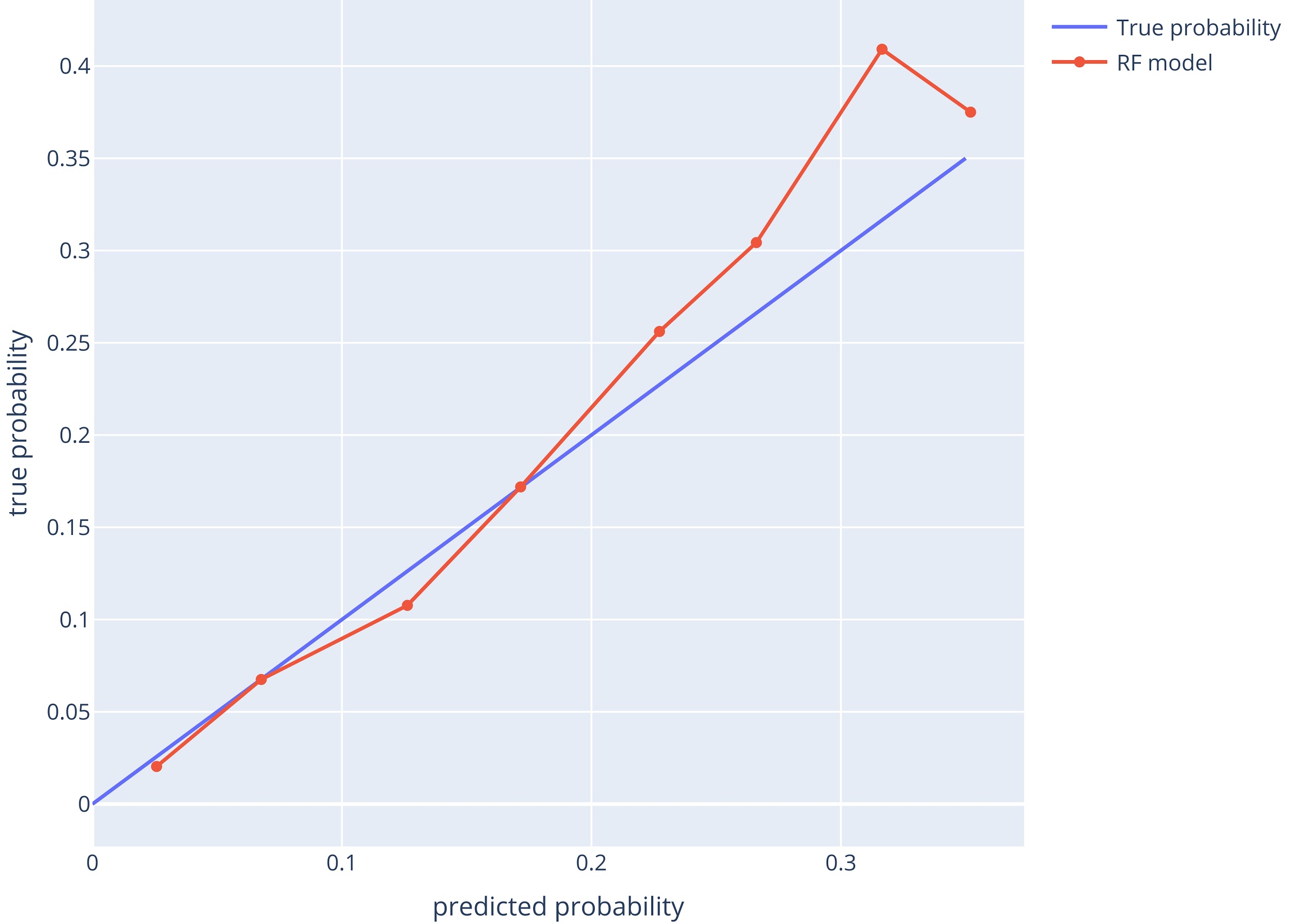

**Appendix Figure 5.**  **Calibration of predicted probabilities in the MPM.**

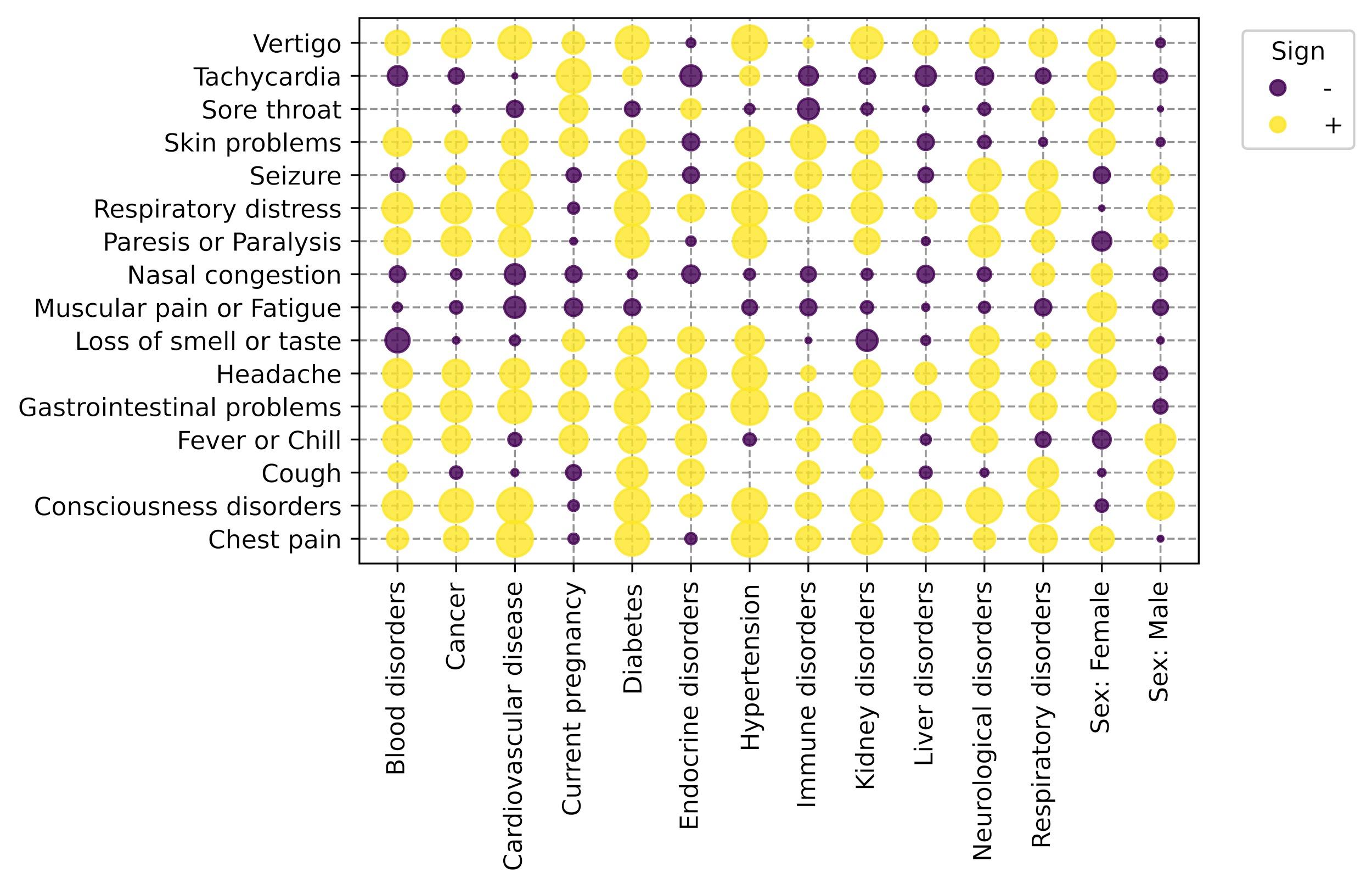

**Appendix Figure 6.** **Chi2 statistics of categorical PMH factors in the SPM.** The larger the radius, the more two parameters are correlated.

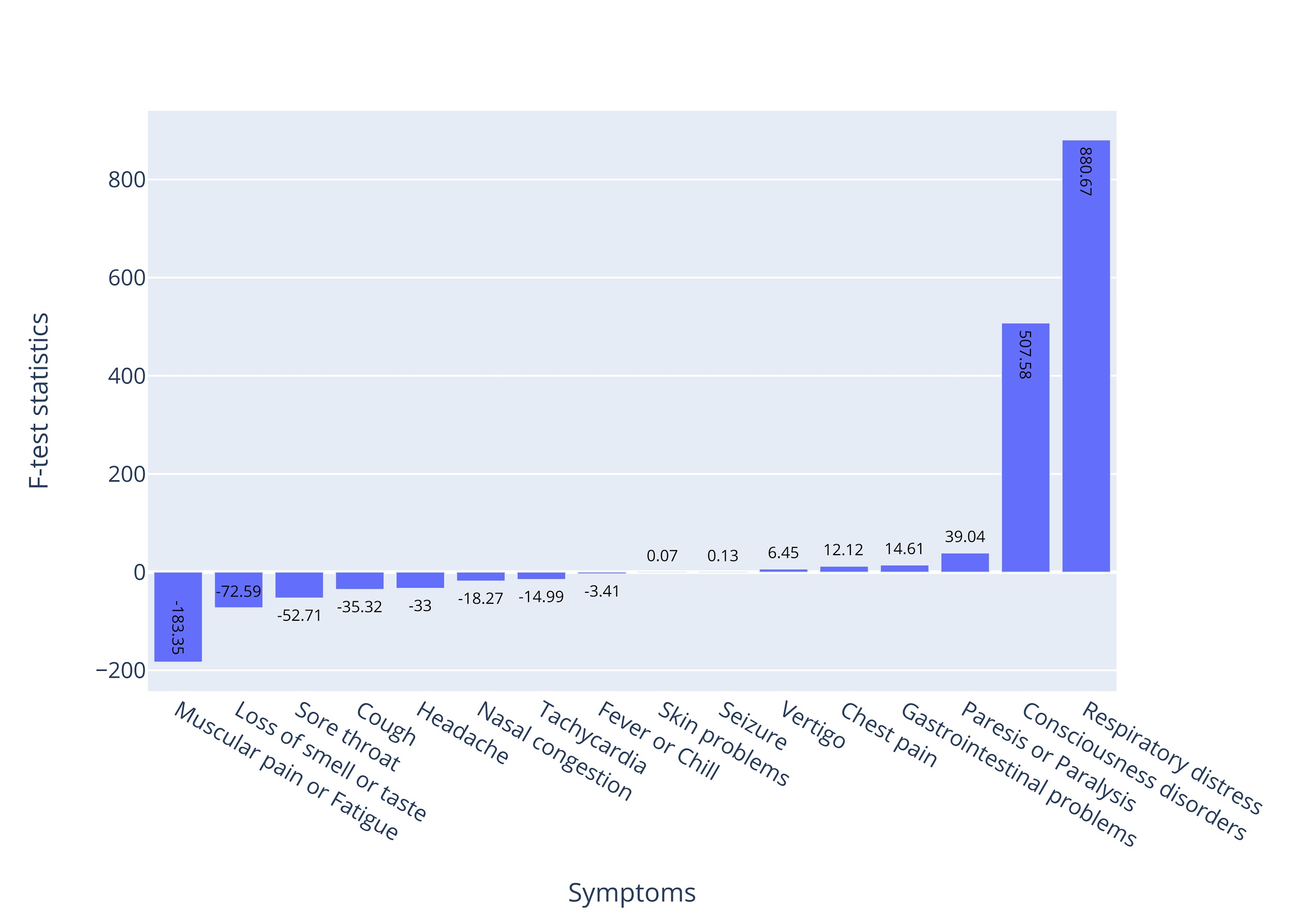

**Appendix Figure 7.** **F-test statistics of age in the SPM.**

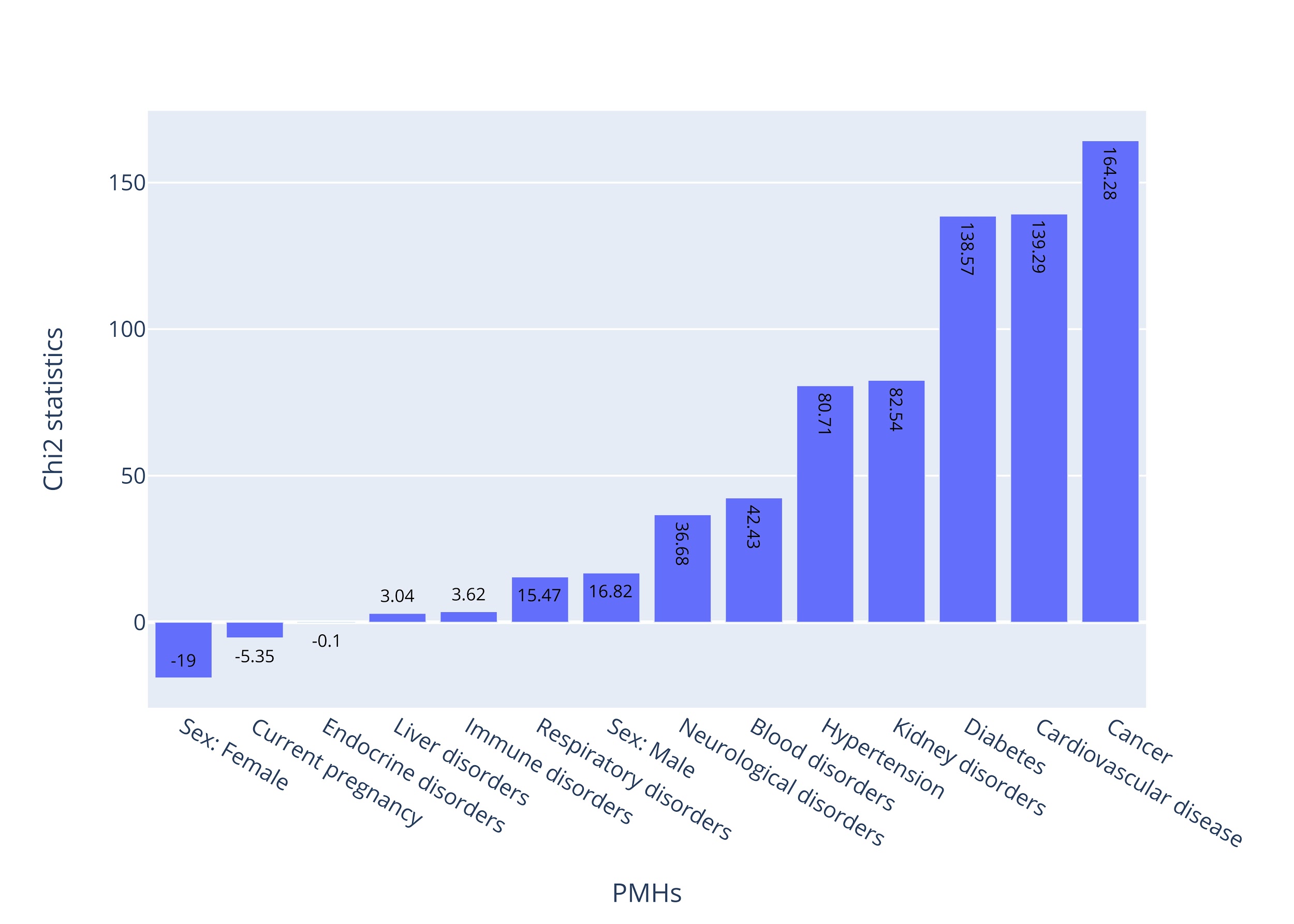

**Appendix Figure 8. Chi2 statistics of categorical past medical history factors in the MPM.**

### Appendix Tables

**Table 1. Comparison of characteristics between COVID-19 patients with and without cough symptom**

| **Cough** | | | | |
| --- | --- | --- | --- | --- |
| **Continue variables** | | | | |
| **Variable** | **Median in not appearance (±IQR)** | **Median appearance (±IQR)** | **F-test statistics** | **F-test p-value** |
| **Age** | 53 (±30) | 51 (±29) | 35.32 | <0.001 |
| **Categorical/Binary variables** | | | | |
| **Variable** | **Count in not appearance (percent in not appearance)** | **Count in appearance (percent in appearance)** | **Chi2 statics** | **Chi2 p-value** |
| **Sex** |  |  |  |  |
| Male | 6,169 (52.48%) | 6,428 (53.59%) | 1.37 | 0.243 |
| Female | 5,585 (47.52%) | 5,567 (46.41%) | 1.54 | 0.214 |
| **Cardiovascular disease** | 1,253 (10.66%) | 1,218 (10.15%) | 1.46 | 0.227 |
| **Diabetes** | 935 (7.95%) | 1,133 (9.45%) | 15.15 | <0.001 |
| **Hypertension** | 992 (8.44%) | 1,012 (8.44%) | 0 | 0.994 |
| **Respiratory disorders** | 229 (1.95%) | 317 (2.64%) | 12.45 | <0.001 |
| **Cancer** | 256 (2.18%) | 221 (1.84%) | 3.33 | 0.068 |
| **Kidney disorders** | 204 (1.74%) | 212 (1.77%) | 0.03 | 0.853 |
| **Neurological disorders** | 137 (1.17%) | 127 (1.06%) | 0.61 | 0.435 |
| **Immune disorders** | 83 (0.71%) | 95 (0.79%) | 0.58 | 0.445 |
| **Blood disorders** | 73 (0.62%) | 79 (0.66%) | 0.13 | 0.718 |
| **Current pregnancy** | 83 (0.71%) | 56 (0.47%) | 5.81 | 0.016 |
| **Liver disorders** | 62 (0.53%) | 57 (0.48%) | 0.32 | 0.569 |
| **Endocrine disorders** | 41 (0.35%) | 56 (0.47%) | 2.03 | 0.155 |
| **Organ or bone marrow Transplant** | 16 (0.14%) | 13 (0.11%) | 0.37 | 0.541 |
| **Psychiatric disorders** | 6 (0.05%) | 13 (0.11%) | 2.44 | 0.118 |
| **Thrombosis** | 7 (0.06%) | 8 (0.07%) | 0.05 | 0.827 |
| **Past COVID-19 infection** | 5 (0.04%) | 5 (0.04%) | 0 | 0.974 |

**Table 2. Comparison of characteristics between COVID-19 patients with and without respiratory distress**

| **Respiratory Distress** | | | | |
| --- | --- | --- | --- | --- |
| **Continue variables** | | | | |
| **Variable** | **Median in not appearance (±IQR)** | **Median appearance (±IQR)** | **F-test statistics** | **F-test p-value** |
| **Age** | 47 (±28) | 57 (±29) | 880.67 | <0.001 |
| **Categorical/Binary variables** | | | | |
| **Variable** | **Count in not appearance (percent in not appearance)** | **Count in appearance (percent in appearance)** | **Chi2 statics** | **Chi2 p-value** |
| **Sex** |  |  |  |  |
| Male | 7,053 (52.61%) | 5,544 (53.61%) | 1.1 | 0.294 |
| Female | 6,354 (47.39%) | 4,798 (46.39%) | 1.24 | 0.265 |
| **Cardiovascular disease** | 1,014 (7.56%) | 1,457 (14.09%) | 238.9 | <0.001 |
| **Diabetes** | 909 (6.78%) | 1,159 (11.21%) | 131.38 | <0.001 |
| **Hypertension** | 855 (6.38%) | 1,149 (11.11%) | 154.98 | <0.001 |
| **Respiratory disorders** | 186 (1.39%) | 360 (3.48%) | 111.31 | <0.001 |
| **Cancer** | 229 (1.71%) | 248 (2.4%) | 13.84 | <0.001 |
| **Kidney disorders** | 190 (1.42%) | 226 (2.19%) | 19.66 | <0.001 |
| **Neurological disorders** | 134 (1.0%) | 130 (1.26%) | 3.48 | 0.062 |
| **Immune disorders** | 90 (0.67%) | 88 (0.85%) | 2.51 | 0.113 |
| **Blood disorders** | 64 (0.48%) | 88 (0.85%) | 12.73 | <0.001 |
| **Current pregnancy** | 88 (0.66%) | 51 (0.49%) | 2.66 | 0.103 |
| **Liver disorders** | 64 (0.48%) | 55 (0.53%) | 0.35 | 0.557 |
| **Endocrine disorders** | 47 (0.35%) | 50 (0.48%) | 2.52 | 0.112 |
| **Organ or bone marrow Transplant** | 16 (0.12%) | 13 (0.13%) | 0.02 | 0.889 |
| **Psychiatric disorders** | 10 (0.07%) | 9 (0.09%) | 0.11 | 0.737 |
| **Thrombosis** | 5 (0.04%) | 10 (0.1%) | 3.26 | 0.071 |
| **Past COVID-19 infection** | 8 (0.06%) | 2 (0.02%) | 2.26 | 0.133 |

**Table 3. Comparison of characteristics between COVID-19 patients with and without muscular pain or fatigue**

| **Muscular Pain or Fatigue** | | | | |
| --- | --- | --- | --- | --- |
| **Continue variables** | | | | |
| **Variable** | **Median in not appearance (±IQR)** | **Median appearance (±IQR)** | **F-test statistics** | **F-test p-value** |
| **Age** | 54 (±29) | 49 (±29) | 183.35 | <0.001 |
| **Categorical/Binary variables** | | | | |
| **Variable** | **Count in not appearance (percent in not appearance)** | **Count in appearance (percent in appearance)** | **Chi2 statics** | **Chi2 p-value** |
| **Sex** |  |  |  |  |
| Male | 7,824 (53.96%) | 4,773 (51.61%) | 5.89 | 0.015 |
| Female | 6,676 (46.04%) | 4,476 (48.39%) | 6.66 | 0.01 |
| **Cardiovascular disease** | 1,649 (11.37%) | 822 (8.89%) | 33.51 | <0.001 |
| **Diabetes** | 1,324 (9.13%) | 744 (8.04%) | 7.66 | 0.006 |
| **Hypertension** | 1,275 (8.79%) | 729 (7.88%) | 5.56 | 0.018 |
| **Respiratory disorders** | 366 (2.52%) | 180 (1.95%) | 8.21 | 0.004 |
| **Cancer** | 297 (2.05%) | 180 (1.95%) | 0.29 | 0.588 |
| **Kidney disorders** | 272 (1.88%) | 144 (1.56%) | 3.28 | 0.07 |
| **Neurological disorders** | 174 (1.2%) | 90 (0.97%) | 2.62 | 0.106 |
| **Immune disorders** | 111 (0.77%) | 67 (0.72%) | 0.13 | 0.721 |
| **Blood disorders** | 101 (0.7%) | 51 (0.55%) | 1.86 | 0.173 |
| **Current pregnancy** | 104 (0.72%) | 35 (0.38%) | 11.08 | 0.001 |
| **Liver disorders** | 77 (0.53%) | 42 (0.45%) | 0.67 | 0.414 |
| **Endocrine disorders** | 59 (0.41%) | 38 (0.41%) | 0 | 0.963 |
| **Organ or bone marrow Transplant** | 22 (0.15%) | 7 (0.08%) | 2.67 | 0.102 |
| **Psychiatric disorders** | 14 (0.1%) | 5 (0.05%) | 1.27 | 0.259 |
| **Thrombosis** | 10 (0.07%) | 5 (0.05%) | 0.2 | 0.656 |
| **Past COVID-19 infection** | 6 (0.04%) | 4 (0.04%) | 0 | 0.945 |

**Table 4. Comparison of characteristics between COVID-19 patients with and without fever or chill**

| **Fever or Chill** | | | | |
| --- | --- | --- | --- | --- |
| **Continue variables** | | | | |
| **Variable** | **Median in not appearance (±IQR)** | **Median appearance (±IQR)** | **F-test statistics** | **F-test p-value** |
| **Age** | 52 (±29) | 51 (±28) | 3.41 | 0.065 |
| **Categorical/Binary variables** | | | | |
| **Variable** | **Count in not appearance (percent in not appearance)** | **Count in appearance (percent in appearance)** | **Chi2 statics** | **Chi2 p-value** |
| **Sex** |  |  |  |  |
| Male | 7,883 (51.88%) | 4,714 (55.12%) | 10.83 | 0.001 |
| Female | 7,313 (48.12%) | 3,839 (44.88%) | 12.23 | <0.001 |
| **Cardiovascular disease** | 1,628 (10.71%) | 843 (9.86%) | 3.86 | 0.049 |
| **Diabetes** | 1,284 (8.45%) | 784 (9.17%) | 3.23 | 0.072 |
| **Hypertension** | 1,321 (8.69%) | 683 (7.99%) | 3.25 | 0.072 |
| **Respiratory disorders** | 354 (2.33%) | 192 (2.24%) | 0.17 | 0.679 |
| **Cancer** | 284 (1.87%) | 193 (2.26%) | 4.09 | 0.043 |
| **Kidney disorders** | 247 (1.63%) | 169 (1.98%) | 3.84 | 0.05 |
| **Neurological disorders** | 158 (1.04%) | 106 (1.24%) | 1.96 | 0.161 |
| **Immune disorders** | 109 (0.72%) | 69 (0.81%) | 0.58 | 0.445 |
| **Blood disorders** | 83 (0.55%) | 69 (0.81%) | 5.8 | 0.016 |
| **Current pregnancy** | 76 (0.5%) | 63 (0.74%) | 5.23 | 0.022 |
| **Liver disorders** | 84 (0.55%) | 35 (0.41%) | 2.25 | 0.134 |
| **Endocrine disorders** | 45 (0.3%) | 52 (0.61%) | 13.03 | <0.001 |
| **Organ or bone marrow Transplant** | 20 (0.13%) | 9 (0.11%) | 0.31 | 0.576 |
| **Psychiatric disorders** | 9 (0.06%) | 10 (0.12%) | 2.28 | 0.131 |
| **Thrombosis** | 10 (0.07%) | 5 (0.06%) | 0.05 | 0.829 |
| **Past COVID-19 infection** | 6 (0.04%) | 4 (0.05%) | 0.07 | 0.793 |

**Table 5. Comparison of characteristics between COVID-19 patients with and without gastrointestinal problems**

| **Gastrointestinal Problems** | | | | |
| --- | --- | --- | --- | --- |
| **Continue variables** | | | | |
| **Variable** | **Median in not appearance (±IQR)** | **Median appearance (±IQR)** | **F-test statistics** | **F-test p-value** |
| **Age** | 51.5 (±28) | 54 (±31) | 14.61 | <0.001 |
| **Categorical/Binary variables** | | | | |
| **Variable** | **Count in not appearance (percent in not appearance)** | **Count in appearance (percent in appearance)** | **Chi2 statics** | **Chi2 p-value** |
| **Sex** |  |  |  |  |
| Male | 11,362 (53.39%) | 1,235 (50.02%) | 4.74 | 0.029 |
| Female | 9,918 (46.61%) | 1,234 (49.98%) | 5.36 | 0.021 |
| **Cardiovascular disease** | 2,097 (9.85%) | 374 (15.15%) | 59.58 | <0.001 |
| **Diabetes** | 1,689 (7.94%) | 379 (15.35%) | 139.63 | <0.001 |
| **Hypertension** | 1,533 (7.2%) | 471 (19.08%) | 369.56 | <0.001 |
| **Respiratory disorders** | 478 (2.25%) | 68 (2.75%) | 2.48 | 0.115 |
| **Cancer** | 399 (1.88%) | 78 (3.16%) | 18.16 | <0.001 |
| **Kidney disorders** | 338 (1.59%) | 78 (3.16%) | 31.16 | <0.001 |
| **Neurological disorders** | 219 (1.03%) | 45 (1.82%) | 12.53 | <0.001 |
| **Immune disorders** | 152 (0.71%) | 26 (1.05%) | 3.39 | 0.066 |
| **Blood disorders** | 129 (0.61%) | 23 (0.93%) | 3.66 | 0.056 |
| **Current pregnancy** | 113 (0.53%) | 26 (1.05%) | 10.3 | 0.001 |
| **Liver disorders** | 95 (0.45%) | 24 (0.97%) | 12.2 | <0.001 |
| **Endocrine disorders** | 82 (0.39%) | 15 (0.61%) | 2.67 | 0.102 |
| **Organ or bone marrow Transplant** | 25 (0.12%) | 4 (0.16%) | 0.36 | 0.549 |
| **Psychiatric disorders** | 14 (0.07%) | 5 (0.2%) | 5.17 | 0.023 |
| **Thrombosis** | 10 (0.05%) | 5 (0.2%) | 8.47 | 0.004 |
| **Past COVID-19 infection** | 9 (0.04%) | 1 (0.04%) | 0 | 0.967 |

**Table 6. Comparison of characteristics between COVID-19 patients with and without headache**

| **Headache** | | | | |
| --- | --- | --- | --- | --- |
| **Continue variables** | | | | |
| **Variable** | **Median in not appearance (±IQR)** | **Median appearance (±IQR)** | **F-test statistics** | **F-test p-value** |
| **Age** | 52 (±29) | 48 (±28) | 33 | <0.001 |
| **Categorical/Binary variables** | | | | |
| **Variable** | **Count in not appearance (percent in not appearance)** | **Count in appearance (percent in appearance)** | **Chi2 statics** | **Chi2 p-value** |
| **Sex** |  |  |  |  |
| Male | 12,050 (53.25%) | 547 (48.84%) | 3.91 | 0.048 |
| Female | 10,579 (46.75%) | 573 (51.16%) | 4.42 | 0.035 |
| **Cardiovascular disease** | 2,324 (10.27%) | 147 (13.12%) | 8.36 | 0.004 |
| **Diabetes** | 1,908 (8.43%) | 160 (14.29%) | 42 | <0.001 |
| **Hypertension** | 1,818 (8.03%) | 186 (16.61%) | 92.95 | <0.001 |
| **Respiratory disorders** | 515 (2.28%) | 31 (2.77%) | 1.12 | 0.289 |
| **Cancer** | 446 (1.97%) | 31 (2.77%) | 3.37 | 0.066 |
| **Kidney disorders** | 390 (1.72%) | 26 (2.32%) | 2.18 | 0.14 |
| **Neurological disorders** | 242 (1.07%) | 22 (1.96%) | 7.69 | 0.006 |
| **Immune disorders** | 169 (0.75%) | 9 (0.8%) | 0.05 | 0.83 |
| **Blood disorders** | 138 (0.61%) | 14 (1.25%) | 6.83 | 0.009 |
| **Current pregnancy** | 129 (0.57%) | 10 (0.89%) | 1.9 | 0.168 |
| **Liver disorders** | 112 (0.49%) | 7 (0.62%) | 0.36 | 0.548 |
| **Endocrine disorders** | 85 (0.38%) | 12 (1.07%) | 12.65 | <0.001 |
| **Organ or bone marrow Transplant** | 25 (0.11%) | 4 (0.36%) | 5.32 | 0.021 |
| **Psychiatric disorders** | 17 (0.08%) | 2 (0.18%) | 1.43 | 0.232 |
| **Thrombosis** | 15 (0.07%) | 0 (0.0%) | 0.74 | 0.389 |
| **Past COVID-19 infection** | 8 (0.04%) | 2 (0.18%) | 5.2 | 0.023 |

**Table 7. Comparison of characteristics between COVID-19 patients with and without chest pain**

| **Chest Pain** | | | | |
| --- | --- | --- | --- | --- |
| **Continue variables** | | | | |
| **Variable** | **Median in not appearance (±IQR)** | **Median appearance (±IQR)** | **F-test statistics** | **F-test p-value** |
| **Age** | 52 (±29) | 55 (±27) | 12.12 | <0.001 |
| **Categorical/Binary variables** | | | | |
| **Variable** | **Count in not appearance (percent in not appearance)** | **Count in appearance (percent in appearance)** | **Chi2 statics** | **Chi2 p-value** |
| **Sex** |  |  |  |  |
| Male | 12,219 (53.12%) | 378 (50.74%) | 0.77 | 0.38 |
| Female | 10,785 (46.88%) | 367 (49.26%) | 0.87 | 0.351 |
| **Cardiovascular disease** | 2,250 (9.78%) | 221 (29.66%) | 274.2 | <0.001 |
| **Diabetes** | 1,932 (8.4%) | 136 (18.26%) | 80.51 | <0.001 |
| **Hypertension** | 1,817 (7.9%) | 187 (25.1%) | 253.06 | <0.001 |
| **Respiratory disorders** | 521 (2.26%) | 25 (3.36%) | 3.74 | 0.053 |
| **Cancer** | 458 (1.99%) | 19 (2.55%) | 1.12 | 0.289 |
| **Kidney disorders** | 389 (1.69%) | 27 (3.62%) | 15.4 | <0.001 |
| **Neurological disorders** | 254 (1.1%) | 10 (1.34%) | 0.37 | 0.544 |
| **Immune disorders** | 170 (0.74%) | 8 (1.07%) | 1.08 | 0.299 |
| **Blood disorders** | 146 (0.63%) | 6 (0.81%) | 0.33 | 0.567 |
| **Current pregnancy** | 136 (0.59%) | 3 (0.4%) | 0.44 | 0.508 |
| **Liver disorders** | 113 (0.49%) | 6 (0.81%) | 1.42 | 0.233 |
| **Endocrine disorders** | 95 (0.41%) | 2 (0.27%) | 0.37 | 0.544 |
| **Organ or bone marrow Transplant** | 27 (0.12%) | 2 (0.27%) | 1.35 | 0.245 |
| **Psychiatric disorders** | 19 (0.08%) | 0 (0.0%) | 0.62 | 0.433 |
| **Thrombosis** | 15 (0.07%) | 0 (0.0%) | 0.49 | 0.486 |
| **Past COVID-19 infection** | 10 (0.04%) | 0 (0.0%) | 0.32 | 0.569 |

**Table 8. Comparison of characteristics between covid-19 patients with and without consciousness disorder**

| **Consciousness Disorders** | | | | |
| --- | --- | --- | --- | --- |
| **Continue variables** | | | | |
| **Variable** | **Median in not appearance (±IQR)** | **Median appearance (±IQR)** | **F-test statistics** | **F-test p-value** |
| **Age** | 51 (±29) | 71 (±22) | 507.58 | <0.001 |
| **Categorical/Binary variables** | | | | |
| **Variable** | **Count in not appearance (percent in not appearance)** | **Count in appearance (percent in appearance)** | **Chi2 statics** | **Chi2 p-value** |
| **Sex** |  |  |  |  |
| Male | 12,194 (52.9%) | 403 (57.74%) | 2.99 | 0.084 |
| Female | 10,857 (47.1%) | 295 (42.26%) | 3.37 | 0.066 |
| **Cardiovascular disease** | 2,273 (9.86%) | 198 (28.37%) | 223 | <0.001 |
| **Diabetes** | 1,912 (8.29%) | 156 (22.35%) | 153.69 | <0.001 |
| **Hypertension** | 1,863 (8.08%) | 141 (20.2%) | 117.91 | <0.001 |
| **Respiratory disorders** | 504 (2.19%) | 42 (6.02%) | 43.24 | <0.001 |
| **Cancer** | 432 (1.87%) | 45 (6.45%) | 70.54 | <0.001 |
| **Kidney disorders** | 382 (1.66%) | 34 (4.87%) | 39.95 | <0.001 |
| **Neurological disorders** | 216 (0.94%) | 48 (6.88%) | 215.02 | <0.001 |
| **Immune disorders** | 170 (0.74%) | 8 (1.15%) | 1.51 | 0.219 |
| **Blood disorders** | 141 (0.61%) | 11 (1.58%) | 9.84 | 0.002 |
| **Current pregnancy** | 138 (0.6%) | 1 (0.14%) | 2.4 | 0.121 |
| **Liver disorders** | 104 (0.45%) | 15 (2.15%) | 38.97 | <0.001 |
| **Endocrine disorders** | 93 (0.4%) | 4 (0.57%) | 0.48 | 0.49 |
| **Organ or bone marrow Transplant** | 29 (0.13%) | 0 (0.0%) | 0.88 | 0.349 |
| **Psychiatric disorders** | 17 (0.07%) | 2 (0.29%) | 3.83 | 0.05 |
| **Thrombosis** | 15 (0.07%) | 0 (0.0%) | 0.45 | 0.5 |
| **Past COVID-19 infection** | 10 (0.04%) | 0 (0.0%) | 0.3 | 0.582 |

**Table 9. Comparison of characteristics between COVID-19 patients with and without loss of smell or taste**

| **Loss of Smell or Taste** | | | | |
| --- | --- | --- | --- | --- |
| **Continue variables** | | | | |
| **Variable** | **Median in not appearance (±IQR)** | **Median appearance (±IQR)** | **F-test statistics** | **F-test p-value** |
| **Age** | 52 (±29) | 42 (±25) | 72.59 | <0.001 |
| **Categorical/Binary variables** | | | | |
| **Variable** | **Count in not appearance (percent in not appearance)** | **Count in appearance (percent in appearance)** | **Chi2 statics** | **Chi2 p-value** |
| **Sex** |  |  |  |  |
| Male | 12,269 (53.14%) | 328 (49.77%) | 1.37 | 0.242 |
| Female | 10,821 (46.86%) | 331 (50.23%) | 1.54 | 0.214 |
| **Cardiovascular disease** | 2,408 (10.43%) | 63 (9.56%) | 0.46 | 0.495 |
| **Diabetes** | 1,995 (8.64%) | 73 (11.08%) | 4.37 | 0.037 |
| **Hypertension** | 1,931 (8.36%) | 73 (11.08%) | 5.59 | 0.018 |
| **Respiratory disorders** | 530 (2.3%) | 16 (2.43%) | 0.05 | 0.825 |
| **Cancer** | 468 (2.03%) | 9 (1.37%) | 1.39 | 0.238 |
| **Kidney disorders** | 405 (1.75%) | 11 (1.67%) | 0.03 | 0.871 |
| **Neurological disorders** | 250 (1.08%) | 14 (2.12%) | 6.25 | 0.012 |
| **Immune disorders** | 175 (0.76%) | 3 (0.46%) | 0.78 | 0.376 |
| **Blood disorders** | 148 (0.64%) | 4 (0.61%) | 0.01 | 0.914 |
| **Current pregnancy** | 134 (0.58%) | 5 (0.76%) | 0.35 | 0.555 |
| **Liver disorders** | 117 (0.51%) | 2 (0.3%) | 0.53 | 0.467 |
| **Endocrine disorders** | 92 (0.4%) | 5 (0.76%) | 2.04 | 0.154 |
| **Organ or bone marrow Transplant** | 29 (0.13%) | 0 (0.0%) | 0.83 | 0.363 |
| **Psychiatric disorders** | 19 (0.08%) | 0 (0.0%) | 0.54 | 0.461 |
| **Thrombosis** | 15 (0.06%) | 0 (0.0%) | 0.43 | 0.513 |
| **Past COVID-19 infection** | 9 (0.04%) | 1 (0.15%) | 1.93 | 0.164 |

**Table 10. Comparison of characteristics between COVID-19 patients with and without vertigo**

| **Vertigo** | | | | |
| --- | --- | --- | --- | --- |
| **Continue variables** | | | | |
| **Variable** | **Median in not appearance (±IQR)** | **Median appearance (±IQR)** | **F-test statistics** | **F-test p-value** |
| **Age** | 52 (±29) | 55 (±29) | 6.45 | 0.011 |
| **Categorical/Binary variables** | | | | |
| **Variable** | **Count in not appearance (percent in not appearance)** | **Count in appearance (percent in appearance)** | **Chi2 statics** | **Chi2 p-value** |
| **Sex** |  |  |  |  |
| Male | 12,353 (53.14%) | 244 (48.7%) | 1.82 | 0.178 |
| Female | 10,895 (46.86%) | 257 (51.3%) | 2.05 | 0.152 |
| **Cardiovascular disease** | 2,367 (10.18%) | 104 (20.76%) | 52.73 | <0.001 |
| **Diabetes** | 1,976 (8.5%) | 92 (18.36%) | 54.8 | <0.001 |
| **Hypertension** | 1,890 (8.13%) | 114 (22.75%) | 124.31 | <0.001 |
| **Respiratory disorders** | 528 (2.27%) | 18 (3.59%) | 3.73 | 0.054 |
| **Cancer** | 458 (1.97%) | 19 (3.79%) | 8.11 | 0.004 |
| **Kidney disorders** | 392 (1.69%) | 24 (4.79%) | 26.98 | <0.001 |
| **Neurological disorders** | 252 (1.08%) | 12 (2.4%) | 7.59 | 0.006 |
| **Immune disorders** | 174 (0.75%) | 4 (0.8%) | 0.02 | 0.898 |
| **Blood disorders** | 147 (0.63%) | 5 (1.0%) | 1.02 | 0.311 |
| **Current pregnancy** | 135 (0.58%) | 4 (0.8%) | 0.4 | 0.529 |
| **Liver disorders** | 115 (0.49%) | 4 (0.8%) | 0.9 | 0.342 |
| **Endocrine disorders** | 96 (0.41%) | 1 (0.2%) | 0.55 | 0.46 |
| **Organ or bone marrow Transplant** | 26 (0.11%) | 3 (0.6%) | 9.52 | 0.002 |
| **Psychiatric disorders** | 19 (0.08%) | 0 (0.0%) | 0.41 | 0.522 |
| **Thrombosis** | 15 (0.06%) | 0 (0.0%) | 0.32 | 0.57 |
| **Past COVID-19 infection** | 10 (0.04%) | 0 (0.0%) | 0.22 | 0.642 |

**Table 11. Comparison of characteristics between COVID-19 patients with and without sore throat**

| **Sore Throat** | | | | |
| --- | --- | --- | --- | --- |
| **Continue variables** | | | | |
| **Variable** | **Median in not appearance (±IQR)** | **Median appearance (±IQR)** | **F-test statistics** | **F-test p-value** |
| **Age** | 52 (±29) | 38 (±19) | 52.71 | <0.001 |
| **Categorical/Binary variables** | | | | |
| **Variable** | **Count in not appearance (percent in not appearance)** | **Count in appearance (percent in appearance)** | **Chi2 statics** | **Chi2 p-value** |
| **Sex** |  |  |  |  |
| Male | 12,522 (53.08%) | 75 (47.77%) | 0.83 | 0.363 |
| Female | 11,070 (46.92%) | 82 (52.23%) | 0.94 | 0.333 |
| **Cardiovascular disease** | 2,466 (10.45%) | 5 (3.18%) | 7.92 | 0.005 |
| **Diabetes** | 2,063 (8.74%) | 5 (3.18%) | 5.54 | 0.019 |
| **Hypertension** | 1,996 (8.46%) | 8 (5.1%) | 2.09 | 0.148 |
| **Respiratory disorders** | 541 (2.29%) | 5 (3.18%) | 0.54 | 0.463 |
| **Cancer** | 476 (2.02%) | 1 (0.64%) | 1.48 | 0.224 |
| **Kidney disorders** | 416 (1.76%) | 0 (0.0%) | 2.77 | 0.096 |
| **Neurological disorders** | 263 (1.11%) | 1 (0.64%) | 0.32 | 0.571 |
| **Immune disorders** | 177 (0.75%) | 1 (0.64%) | 0.03 | 0.87 |
| **Blood disorders** | 151 (0.64%) | 1 (0.64%) | 0 | 0.996 |
| **Current pregnancy** | 136 (0.58%) | 3 (1.91%) | 4.74 | 0.029 |
| **Liver disorders** | 119 (0.5%) | 0 (0.0%) | 0.79 | 0.374 |
| **Endocrine disorders** | 96 (0.41%) | 1 (0.64%) | 0.2 | 0.653 |
| **Organ or bone marrow Transplant** | 28 (0.12%) | 1 (0.64%) | 3.43 | 0.064 |
| **Psychiatric disorders** | 19 (0.08%) | 0 (0.0%) | 0.13 | 0.722 |
| **Thrombosis** | 15 (0.06%) | 0 (0.0%) | 0.1 | 0.752 |
| **Past COVID-19 infection** | 9 (0.04%) | 1 (0.64%) | 13.28 | <0.001 |

**Table 12. Comparison of characteristics between COVID-19 patients with and without paresis or paralysis.**

| **Paresis or Paralysis** | | | | |
| --- | --- | --- | --- | --- |
| **Continue variables** | | | | |
| **Variable** | **Median in not appearance (±IQR)** | **Median appearance (±IQR)** | **F-test statistics** | **F-test p-value** |
| **Age** | 52 (±29) | 67 (±21) | 39.04 | <0.001 |
| **Categorical/Binary variables** | | | | |
| **Variable** | **Count in not appearance (percent in not appearance)** | **Count in appearance (percent in appearance)** | **Chi2 statics** | **Chi2 p-value** |
| **Sex** |  |  |  |  |
| Male | 12,531 (53.03%) | 66 (54.55%) | 0.05 | 0.82 |
| Female | 11,097 (46.97%) | 55 (45.45%) | 0.06 | 0.809 |
| **Cardiovascular disease** | 2,442 (10.34%) | 29 (23.97%) | 21.5 | <0.001 |
| **Diabetes** | 2,034 (8.61%) | 34 (28.1%) | 52.52 | <0.001 |
| **Hypertension** | 1,969 (8.33%) | 35 (28.93%) | 60.5 | <0.001 |
| **Respiratory disorders** | 542 (2.29%) | 4 (3.31%) | 0.54 | 0.464 |
| **Cancer** | 470 (1.99%) | 7 (5.79%) | 8.64 | 0.003 |
| **Kidney disorders** | 412 (1.74%) | 4 (3.31%) | 1.68 | 0.195 |
| **Neurological disorders** | 257 (1.09%) | 7 (5.79%) | 23.9 | <0.001 |
| **Immune disorders** | 177 (0.75%) | 1 (0.83%) | 0.01 | 0.922 |
| **Blood disorders** | 150 (0.63%) | 2 (1.65%) | 1.95 | 0.163 |
| **Current pregnancy** | 139 (0.59%) | 0 (0.0%) | 0.71 | 0.399 |
| **Liver disorders** | 119 (0.5%) | 0 (0.0%) | 0.61 | 0.435 |
| **Endocrine disorders** | 97 (0.41%) | 0 (0.0%) | 0.5 | 0.481 |
| **Organ or bone marrow Transplant** | 29 (0.12%) | 0 (0.0%) | 0.15 | 0.7 |
| **Psychiatric disorders** | 19 (0.08%) | 0 (0.0%) | 0.1 | 0.755 |
| **Thrombosis** | 15 (0.06%) | 0 (0.0%) | 0.08 | 0.782 |
| **Past COVID-19 infection** | 10 (0.04%) | 0 (0.0%) | 0.05 | 0.821 |

**Table 13. Comparison of characteristics between COVID-19 patients with and without seizure.**

| **Seizure** | | | | |
| --- | --- | --- | --- | --- |
| **Continue variables** | | | | |
| **Variable** | **Median in not appearance (±IQR)** | **Median appearance (±IQR)** | **F-test statistics** | **F-test p-value** |
| **Age** | 52 (±29) | 55 (±21.5) | 0.13 | 0.72 |
| **Categorical/Binary variables** | | | | |
| **Variable** | **Count in not appearance (percent in not appearance)** | **Count in appearance (percent in appearance)** | **Chi2 statics** | **Chi2 p-value** |
| **Sex** |  |  |  |  |
| Male | 12,577 (53.04%) | 20 (57.14%) | 0.11 | 0.739 |
| Female | 11,137 (46.96%) | 15 (42.86%) | 0.13 | 0.723 |
| **Cardiovascular disease** | 2,461 (10.38%) | 10 (28.57%) | 11.12 | 0.001 |
| **Diabetes** | 2,060 (8.69%) | 8 (22.86%) | 8.06 | 0.005 |
| **Hypertension** | 1,999 (8.43%) | 5 (14.29%) | 1.42 | 0.233 |
| **Respiratory disorders** | 543 (2.29%) | 3 (8.57%) | 6 | 0.014 |
| **Cancer** | 476 (2.01%) | 1 (2.86%) | 0.13 | 0.723 |
| **Kidney disorders** | 413 (1.74%) | 3 (8.57%) | 9.31 | 0.002 |
| **Neurological disorders** | 259 (1.09%) | 5 (14.29%) | 54.73 | <0.001 |
| **Immune disorders** | 177 (0.75%) | 1 (2.86%) | 2.08 | 0.149 |
| **Blood disorders** | 152 (0.64%) | 0 (0.0%) | 0.22 | 0.636 |
| **Current pregnancy** | 139 (0.59%) | 0 (0.0%) | 0.21 | 0.651 |
| **Liver disorders** | 119 (0.5%) | 0 (0.0%) | 0.18 | 0.675 |
| **Endocrine disorders** | 97 (0.41%) | 0 (0.0%) | 0.14 | 0.705 |
| **Organ or bone marrow Transplant** | 29 (0.12%) | 0 (0.0%) | 0.04 | 0.836 |
| **Psychiatric disorders** | 19 (0.08%) | 0 (0.0%) | 0.03 | 0.867 |
| **Thrombosis** | 15 (0.06%) | 0 (0.0%) | 0.02 | 0.882 |
| **Past COVID-19 infection** | 10 (0.04%) | 0 (0.0%) | 0.01 | 0.903 |

**Table 14. Comparison of characteristics between COVID-19 patients with and without Skin Problems.**

| **Skin Problems** | | | | |
| --- | --- | --- | --- | --- |
| **Continue variables** | | | | |
| **Variable** | **Median in not appearance (±IQR)** | **Median appearance (±IQR)** | **F-test statistics** | **F-test p-value** |
| **Age** | 52 (±29) | 57 (±35.75) | 0.07 | 0.79 |
| **Categorical/Binary variables** | | | | |
| **Variable** | **Count in not appearance (percent in not appearance)** | **Count in appearance (percent in appearance)** | **Chi2 statics** | **Chi2 p-value** |
| **Sex** |  |  |  |  |
| Male | 12,588 (53.06%) | 9 (34.62%) | 1.67 | 0.197 |
| Female | 11,135 (46.94%) | 17 (65.38%) | 1.88 | 0.17 |
| **Cardiovascular disease** | 2,466 (10.39%) | 5 (19.23%) | 1.95 | 0.163 |
| **Diabetes** | 2,064 (8.7%) | 4 (15.38%) | 1.33 | 0.248 |
| **Hypertension** | 1,998 (8.42%) | 6 (23.08%) | 6.61 | 0.01 |
| **Respiratory disorders** | 546 (2.3%) | 0 (0.0%) | 0.6 | 0.439 |
| **Cancer** | 476 (2.01%) | 1 (3.85%) | 0.44 | 0.508 |
| **Kidney disorders** | 415 (1.75%) | 1 (3.85%) | 0.65 | 0.419 |
| **Neurological disorders** | 264 (1.11%) | 0 (0.0%) | 0.29 | 0.591 |
| **Immune disorders** | 173 (0.73%) | 5 (19.23%) | 118.61 | <0.001 |
| **Blood disorders** | 151 (0.64%) | 1 (3.85%) | 4.18 | 0.041 |
| **Current pregnancy** | 138 (0.58%) | 1 (3.85%) | 4.73 | 0.03 |
| **Liver disorders** | 119 (0.5%) | 0 (0.0%) | 0.13 | 0.718 |
| **Endocrine disorders** | 97 (0.41%) | 0 (0.0%) | 0.11 | 0.744 |
| **Organ or bone marrow Transplant** | 29 (0.12%) | 0 (0.0%) | 0.03 | 0.859 |
| **Psychiatric disorders** | 19 (0.08%) | 0 (0.0%) | 0.02 | 0.885 |
| **Thrombosis** | 15 (0.06%) | 0 (0.0%) | 0.02 | 0.898 |
| **Past COVID-19 infection** | 10 (0.04%) | 0 (0.0%) | 0.01 | 0.917 |

**Table 15. Comparison of characteristics between COVID-19 patients with and without nasal congestion**

| **Nasal Congestion** | | | | |
| --- | --- | --- | --- | --- |
| **Continue variables** | | | | |
| **Variable** | **Median in not appearance (±IQR)** | **Median appearance (±IQR)** | **F-test statistics** | **F-test p-value** |
| **Age** | 52 (±29) | 35 (±6.75) | 18.27 | <0.001 |
| **Categorical/Binary variables** | | | | |
| **Variable** | **Count in not appearance (percent in not appearance)** | **Count in appearance (percent in appearance)** | **Chi2 statics** | **Chi2 p-value** |
| **Sex** |  |  |  |  |
| Male | 12,587 (53.05%) | 10 (45.45%) | 0.24 | 0.625 |
| Female | 11,140 (46.95%) | 12 (54.55%) | 0.27 | 0.603 |
| **Cardiovascular disease** | 2,469 (10.41%) | 2 (9.09%) | 0.04 | 0.848 |
| **Diabetes** | 2,068 (8.72%) | 0 (0.0%) | 1.92 | 0.166 |
| **Hypertension** | 2,003 (8.44%) | 1 (4.55%) | 0.4 | 0.529 |
| **Respiratory disorders** | 545 (2.3%) | 1 (4.55%) | 0.48 | 0.487 |
| **Cancer** | 477 (2.01%) | 0 (0.0%) | 0.44 | 0.506 |
| **Kidney disorders** | 416 (1.75%) | 0 (0.0%) | 0.39 | 0.535 |
| **Neurological disorders** | 264 (1.11%) | 0 (0.0%) | 0.24 | 0.621 |
| **Immune disorders** | 178 (0.75%) | 0 (0.0%) | 0.17 | 0.685 |
| **Blood disorders** | 152 (0.64%) | 0 (0.0%) | 0.14 | 0.707 |
| **Current pregnancy** | 139 (0.59%) | 0 (0.0%) | 0.13 | 0.72 |
| **Liver disorders** | 119 (0.5%) | 0 (0.0%) | 0.11 | 0.74 |
| **Endocrine disorders** | 97 (0.41%) | 0 (0.0%) | 0.09 | 0.764 |
| **Organ or bone marrow Transplant** | 28 (0.12%) | 1 (4.55%) | 35.28 | <0.001 |
| **Psychiatric disorders** | 19 (0.08%) | 0 (0.0%) | 0.02 | 0.894 |
| **Thrombosis** | 15 (0.06%) | 0 (0.0%) | 0.01 | 0.906 |
| **Past COVID-19 infection** | 10 (0.04%) | 0 (0.0%) | 0.01 | 0.923 |

**Table 16. Comparison of characteristics between COVID-19 patients with and without tachycardia**

| **Tachycardia** | | | | |
| --- | --- | --- | --- | --- |
| **Continue variables** | | | | |
| **Variable** | **Median in not appearance (±IQR)** | **Median appearance (±IQR)** | **F-test statistics** | **F-test p-value** |
| **Age** | 52 (±29) | 26.5 (±5.75) | 15 | <0.001 |
| **Categorical/Binary variables** | | | | |
| **Variable** | **Count in not appearance (percent in not appearance)** | **Count in appearance (percent in appearance)** | **Chi2 statics** | **Chi2 p-value** |
| **Sex** |  |  |  |  |
| Male | 12,597 (53.06%) | 0 (0.0%) | 4.24 | 0.039 |
| Female | 11,144 (46.94%) | 8 (100.0%) | 4.79 | 0.029 |
| **Cardiovascular disease** | 2,471 (10.41%) | 0 (0.0%) | 0.83 | 0.362 |
| **Diabetes** | 2,067 (8.71%) | 1 (12.5%) | 0.13 | 0.716 |
| **Hypertension** | 2,003 (8.44%) | 1 (12.5%) | 0.16 | 0.692 |
| **Respiratory disorders** | 546 (2.3%) | 0 (0.0%) | 0.18 | 0.668 |
| **Cancer** | 477 (2.01%) | 0 (0.0%) | 0.16 | 0.688 |
| **Kidney disorders** | 416 (1.75%) | 0 (0.0%) | 0.14 | 0.708 |
| **Neurological disorders** | 264 (1.11%) | 0 (0.0%) | 0.09 | 0.766 |
| **Immune disorders** | 178 (0.75%) | 0 (0.0%) | 0.06 | 0.807 |
| **Blood disorders** | 152 (0.64%) | 0 (0.0%) | 0.05 | 0.821 |
| **Current pregnancy** | 137 (0.58%) | 2 (25.0%) | 81.5 | <0.001 |
| **Liver disorders** | 119 (0.5%) | 0 (0.0%) | 0.04 | 0.841 |
| **Endocrine disorders** | 97 (0.41%) | 0 (0.0%) | 0.03 | 0.857 |
| **Organ or bone marrow Transplant** | 29 (0.12%) | 0 (0.0%) | 0.01 | 0.921 |
| **Psychiatric disorders** | 19 (0.08%) | 0 (0.0%) | 0.01 | 0.936 |
| **Thrombosis** | 15 (0.06%) | 0 (0.0%) | 0.01 | 0.943 |
| **Past COVID-19 infection** | 10 (0.04%) | 0 (0.0%) | 0 | 0.954 |
